# Multi-phase, multi-ethnic GWAS uncovers putative loci in predisposition to human sprint performance, health and disease

**DOI:** 10.1101/2023.12.08.23299720

**Authors:** Guan Wang, Noriyuki Fuku, Eri Miyamoto-Mikami, Masashi Tanaka, Motohiko Miyachi, Haruka Murakami, Yu-Ching Cheng, Braxton D. Mitchell, Errol Morrison, Krista G. Austin, Ildus I. Ahmetov, Sportgene Research Group, Edward V. Generozov, Maxim L. Filipenko, Andrei A. Gilep, Valentina Gineviciene, Colin N. Moran, Tomas Venckunas, Pawel Cieszczyk, Wim Derave, Ioannis Papadimitriou, Fleur C. Garton, Kathryn North, Sandosh Padmanabhan, Yannis P. Pitsiladis

## Abstract

The genetic underpinnings of elite sprint performance remain largely elusive. For the first time, we uncovered rs10196189 (*GALNT13*) in the cross-ancestry, genome-wide analysis of elite sprint and power-oriented athletes and their controls from Jamaica, the USA, and Japan, and replicated this finding in two independent cohorts of elite European athletes (meta-analysis *P* < 5E-08). We identified statistically significant and borderline associations for cross-ancestry and ancestry specific loci in *GALNT13*, *BOP1*, *HSF1*, *STXBP2 GRM7*, *MPRIP*, *ZFYVE28, CERS4*, and *ADAMTS18*, predominantly expressed in the nervous and hematopoietic systems. Further, we revealed thirty-six previously uncharacterized genes associated with host defence, leukocyte migration, and cellular responses to interferon-gamma and unveiled (reprioritized) four genes, *UQCRFS1*, *PTPN6*, *RALY* and *ZMYM4,* responsible for aging, neurological conditions, and blood disorders from the elite athletic performance cohorts. Our results provide new biological insights into elite sprint performance and offer clues to the potential molecular mechanisms interlinking and operating in elite athletic performance and human health and disease.

## Introduction

Over the past two decades, extensive efforts have been made to uncover the genetic basis of elite athletic performance. In a series of updates of the gene map for human performance and fitness-related phenotypes between 2000 and 2015, over 200 autosomal genes and quantitative trait loci have been reported, often lacking replications^1–14^. Genetic studies of human performance continue to produce further results in recent years^15,16^, however, key genes and gene regulatory networks remain largely elusive. The reason behind this knowledge scarcity is multi-faceted, in line with our progressive understanding of the field of human genetics. From the predominant use of a candidate gene approach to applications of a genome-wide approach^17,18^, from the primary focus on European populations to the investigation of genetic diversity across ancestral groups^19–21^, from low to increased statistical power in sample size^22^ and refined case definition^23^, the former has historically contributed to the lack of true genetic associations unveiled so far, while the latter now propels research to tackle complex phenotypes more effectively.

Here, we set out to conduct a multi-phase, cross-ancestry genome-wide association study (GWAS), comprised of elite sprint and power-oriented athletes of West African and East Asian ancestry. These athletes competed in major sprinting and jumping events and represent the tail-end of elite sprinting ability by breaking world records or being Olympic medallists. The extreme phenotype being the top slice of the ability spectrum circumvents the need for a typical GWAS sample size of hundreds of thousands or more. We expect to uncover common genetic variants of moderate to large effect size in the current samples, as it was the case for the landmark GWAS of age-related macular degeneration of a comparable sample size^24^. For the first time, the unique cohorts allowed us, with greater confidence, to explore cross-ancestry and ancestry-specific genetic variations across the genome and their regulatory functions in association with elite sprint and power-oriented athletic status. In addition, we hypothesize that the genetic architecture underlying elite sprinting ability would have significant implications for future studies of human health and disease sharing common biological pathways with elite athletic status.

## Results

### GWAS of elite Jamaican, African-American and Japanese athletes

In the discovery phase, we conducted GWAS using Illumina HumanOmni1-Quad BeadChip (1,134,514 markers) and HumanOmniExpress Beadchip (730,525 markers) in three athlete-control cohorts comprising Jamaicans (Jam; 95 athletes and 102 controls), African-Americans (A-A; 108 athletes and 397 controls) and Japanese (Jpn; 54 athletes and 118 controls). All athletes were specialist sprint and power-oriented athletes with ancestry-matched controls. Standard and cohort-specific quality controls (QCs; Methods) resulted in 609,801, 637,991 and 541,179 markers retained in Jam (88 athletes and 87 controls), A-A (79 athletes and 391 controls) and Jpn (54 athletes and 116 controls) cohorts. Seventeen, seven and twenty-one markers met a suggestive threshold of *P* = 5E-05 in these cohorts, respectively. No variant exceeded the genome-wide significance threshold of *P* = 5E-08.

### Genotype imputation and meta-analyses

Genotype imputation using both IMPUTE2^25^ and Sanger Imputation Server^26^ was carried out for each cohort. The IMPUTE2 pipeline identified 9,726,027 (Jam), 9,640,471 (A-A), and 6,444,519 (Jpn) markers, and the Sanger Imputation Server revealed 7,700,522 (Jam), 7,592,681 (A-A), and 4,975,382 (Jpn) markers (see Supplementary Fig. S1 and S2 for the GWAS and imputation QQ and Manhattan plots). The specific imputation pipelines, post-imputation QCs and association tests adopted are given in Methods.

We then performed separate meta-analyses of the three cohorts following each imputation pipeline. The genomic inflation factor was close to 1 (0.9995, IMPUTE2; and 1.004, Sanger; Supplementary Fig. S3). The meta-analysis results comprised 11,038,143 (IMPUTE2) and 8,787,208 (Sanger) genetic variants, among which 52 (IMPUTE2) and 98 (Sanger) attained the suggestive significance cut-off of *P* = 1E-05 (Figure 1). Notably, the most significant signal of combined effects of all cohorts comes from the imputed variant rs113303758 located in the intron of *GALNT13* on chromosome 2: minor allele frequency (MAF) ranges from 0.06 – 0.35 and the combined odds ratio (OR) is 2.52 for the C-allele (*P*_IMPUTE2_ = 2.75E-07 GC-corrected, *I*^2^ = 0%, *P*_het_ = 0.63; and *P*_Sanger_ = 3.16E-07 GC-corrected, *I*^2^ = 0%, *P*_het_ = 0.64). It is also tightly linked with the GWAS array variant rs10196189 (*r*^2^ = 0.97, *D*’ = 1; OR_IMPUTE2_ = 2.46, *P*_IMPUTE2_ = 6.00E-07 GC-corrected, *I*^2^ = 0%, *P*_het_ = 0.70, and OR_Sanger_ = 2.48, *P*_Sanger_ = 5.45E-07 GC-corrected, *I*^2^ = 0%, *P*_het_ = 0.67, for the G-allele). The association results of rs113303758 and rs10196189 in individual cohorts following Sanger imputation are summarised in Table 1. On a cohort-specific level, the T-allele of rs117143557 (nearest gene: *MPRIP*, and is a distal enhancer-like signature given ENCODE^27^ classification; MAF: 0.06) on chromosome 17 in Jpn had an OR of 17.05 (*P*_Sanger_ = 7.63E-08 GC-corrected), approaching the GWAS significance level of *P* = 5E-08 (Figure 1). In addition, the Sanger imputation results, outperforming IMPUTE2, were fed into the gene-based analyses below.

**Figure 1.**
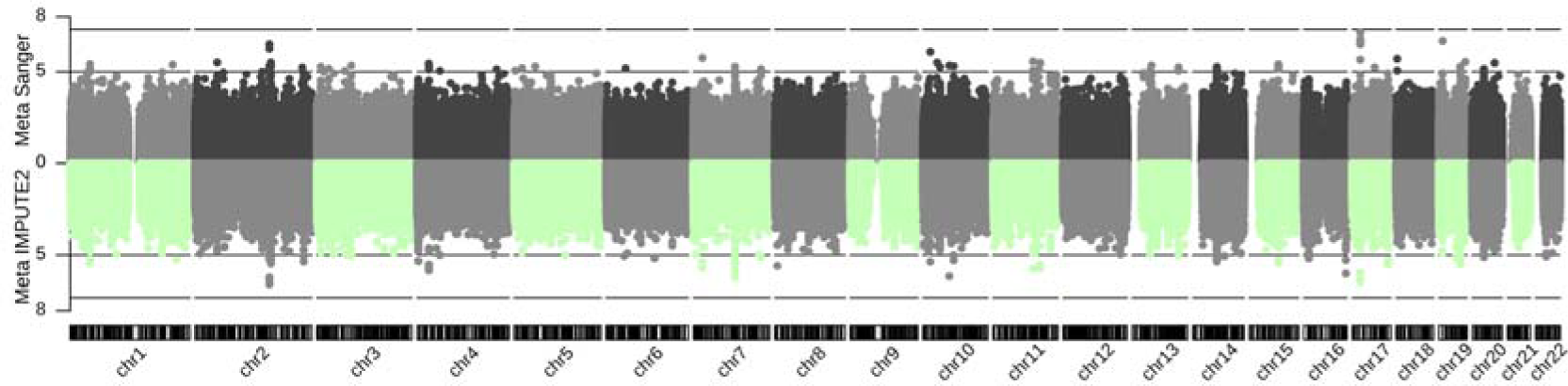
Genetic associations following imputation (IMPUTE2 and Sanger Imputation Server) and meta-analysis of Jam, A-A and Jpn cohorts with athletic prowess (sprinting) across 22 autosomes. Dotted line: suggestive significance cut-off of *P* = 1E-05; dashed line: genome-wide significance cut-off of *P* = 5E-08.

**Table 1.**
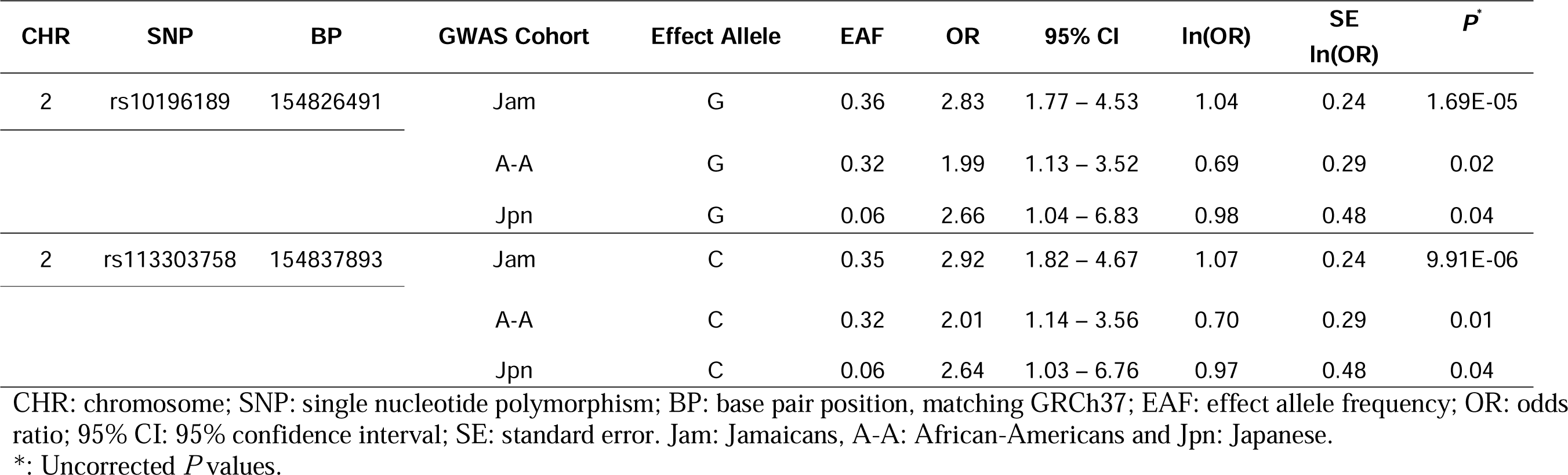
Association results of *GALNT13* rs10196189 and rs113303758 in individual GWAS cohorts following Sanger imputation.

| CHR | SNP | BP | GWAS Cohort | Effect Allele | EA | OR | 95% CI | ln(OR) | SE<br>ln(OR) | *P* * |
| --- | --- | --- | --- | --- | --- | --- | --- | --- | --- | --- |
| 2 | rs10196189 | 154826491 | Jam | G | 0.36 | 2.83 | 1.77 – 4.53 | 1.04 | 0.24 | 1.69E-05 |
|  |  |  | A-A | G | 0.32 | 1.99 | 1.13 – 3.52 | 0.69 | 0.29 | 0.02 |
|  |  |  | Jpn | G | 0.06 | 2.66 | 1.04 – 6.83 | 0.98 | 0.48 | 0.04 |
| 2 | rs113303758 | 154837893 | Jam | C | 0.35 | 2.92 | 1.82 – 4.67 | 1.07 | 0.24 | 9.91E-06 |
|  |  |  | A-A | C | 0.32 | 2.01 | 1.14 – 3.56 | 0.70 | 0.29 | 0.01 |
|  |  |  | Jpn | C | 0.06 | 2.64 | 1.03 – 6.76 | 0.97 | 0.48 | 0.04 |

CHR: chromosome; SNP: single nucleotide polymorphism; BP: base pair position, matching GRCh37; EA: effect allele frequency; OR: odds ratio; 95% CI: 95% confidence interval; SE: standard error. Jam: Jamaicans, A-A: African-Americans and Jpn: Japanese.
\*: Uncorrected *P* values.

### Replication of the GWAS array hit rs10196189 in athletes of European descent

In the replication phase, we took the top hit rs10196189 (in almost perfect linkage disequilibrium, LD, with rs113303758) and independently replicated its association with sprinting ability in two elite European athlete-control cohorts. They comprised 234 power-oriented athletes, 451 endurance-oriented athletes and 1,525 controls from Belarus, Lithuania and Russia (sprint OR_additive_ = 1.53, 95% CI = 1.17 – 2.00, *P* = 2.00E-03; OR_G-dominant_ = 1.61, 95% CI = 1.19 – 2.18, *P* = 1.80E-03, Table 2); and 171 sprint athletes, 252 endurance-oriented athletes and 595 controls from Australia, Belgium, Greece and Poland (sprint OR_additive_ = 1.45, 95% CI = 1.05 – 2.00, *P* = 2.40E-02; OR_A-dominant_ = 0.28, 95% CI = 0.10 – 0.75, *P* = 7.00E-03; Table 2). Details on the replication cohorts for their geographical regions and genotype distribution within each cohort are provided in Supplementary Table S1 and S2. Moreover, meta-analysis of the Sanger imputation and the replication results for rs10196189 revealed a significant association crossing *P* = 5E-08 (OR_additive_= 1.71, *P* = 2.28E-09; *I*^2^ = 41.7%, *P*_het_ = 0.16; and when the sample-size weighted meta-analysis was performed: *P* = 2.13E-09; *I*^2^ = 47.8%, *P*_het_ = 0.11; Table 3).

**Table 2.** Replication results of *GALNT13* rs10196189 in two European cohorts.

| Replication Cohort | Sub Group | Effect Allele*; EAF | Additive Model |  | Dominant Model |  |  |
| --- | --- | --- | --- | --- | --- | --- | --- |
| | | | OR<sup>\$</sup> (95% CI) | *P*<sup>&</sup> | Dominant Allele | OR<sup>¶</sup> (95% CI) | *P*<sup>&</sup> |
| Cohort 1:<br>Belarus, Lithuania, and Russia | Sprinters and Jumpers | (G); 0.14 | 1.23<br>(0.85 – 1.78) | 0.27 | G | 1.32<br>(0.88 – 1.97) | 0.17 |
|  | Other sprint/power oriented | G; 0.17 | 1.53<br>(1.17 – 2.00) | 2.00E-03 | G | 1.61<br>(1.19 – 2.18) | 1.80E-03 |
|  | Endurance oriented | (G); 0.13 | 1.10<br>(0.88 – 1.38) | 0.42 | A | 0.74<br>(0.32 – 1.68) | 0.47 |
| Cohort 2:<br>Australia, Belgium, Greece, and Poland | Sprinters and Jumpers | G; 0.19 | 1.45<br>(1.05 – 2.00) | 2.40E-02 | A | 0.28<br>(0.10 – 0.75) | 7.00E-03 |
|  | Other sprint/power oriented | (G); 0.14 | 1.02<br>(0.71 – 1.46) | 0.92 | A | 1.13<br>(0.24 – 5.38) | 0.88 |
|  | Endurance oriented | (A); 0.88 | 1.23<br>(0.89 – 1.70) | 0.20 | G | 0.70<br>(0.49 – 1.01) | 5.50E-02 |

EAF: effect allele frequency; OR: odds ratio; 95% CI: 95% confidence interval.
\*: The allele whose frequency is higher in the athletes relative to the controls. It bears no meaning where the test results were non-significant (in brackets).
\$: Odds ratios are with respect to the effect allele.
¶: Odds ratios are with respect to the dominant allele.
&: Uncorrected *P* values.

**Table 3.** Meta-analysis of association results for *GALNT13* rs10196189 across the Sanger imputation-GWAS and the two European replication cohorts exceeding the genome-wide significance of *P* = 5E-08.

| SNP | Effect Allele | OR | 95% CI | $P_{\text{combined}}$ | Direction | $I^2$ | $P_{\text{het}}$ |
| --- | --- | --- | --- | --- | --- | --- | --- |
| rs10196189 | G | 1.71 | 1.43 – 2.04 | 2.28E-09 | +++++ | 41.7% | 0.14 |

When sample size weighted meta-analysis performed, $P_{\text{combined}} = 2.13\text{E-}09$ , $I^2 = 47.8\%$ , $P_{\text{het}} = 0.11$ .
SNP: single nucleotide polymorphism; OR: odds ratio; 95% CI: 95% confidence interval; $P_{\text{combined}}$ : combined fixed-effects meta-analysis $P$ value based on the raw additive association $P$ values from each cohort; $I^2$ : heterogeneity index (0-100%); $P_{\text{het}}$ : $P$ value for heterogeneity.

### Gene- and gene-set enrichment analyses

In the gene-analysis phase, we were able to identify 18,416, 18,414, and 18,317 genes containing at least one SNP in Jam, A-A and Jpn, respectively (Methods). Following multiple testing correction, enrichments were observed for *BOP1* (*P*_multi_ = 6.52E-07, and *P*_snpwise_mean_ = 8.35E-07) and *HSF1* (*P*_multi_ = 1.71E-06, and *P*_snpwise_mean_ = 1.67E-06) in Jam, *GRM7* (*P*_multi_ = 1.28E-06) in A-A, and *MPRIP* (*P*_snpwise_top1_ = 1.63E-06) in Jpn. Gene-level meta-analysis of the three cohorts revealed statistically significant enrichment for *ZFYVE28* (*P*_multi_ = 2.01E-06) and a borderline enrichment finding for *ADAMTS18* (*P*_multi_ = 3.13E-06). Competitive gene-set enrichment analysis on the meta-analysis results identified 199 genes significantly enriched in interferon-gamma response (one of the 50 gene sets available from the MSigDB Hallmark Collection^28^; *P* = 7.60E-04 after multiple testing correction); but not in any of the 2,922 Canonical pathways of the MSigDB C2 Collection.

### Gene-based heritability

Gene-based heritability estimation yielded an estimated heritability of 0.20 (SD: 0.09) for *MPRIP* in Jpn on the observed scale, and 0.13 (SD: 0.06) on the liability scale for a prevalence of 1% (test statistic=20.87, *P*_permutation_ = 2.05E-06 following multiple testing correction). This significant finding is concordant with that of the gene enrichment analysis of Jpn reported above. Furthermore, the top gene unveiled in Jam is *STXBP2*, showing the estimated heritability of 0.13 (SD:0.07) and 0.07 (SD: 0.04) on the observed and liability scales, respectively (test statistic = 17.83, *P*_permutation_ = 6.91E-06). For the next two top genes *BOP1* and *HSF1*, the estimated heritability on the observed scale is 0.10 (SD: 0.07) and 0.09 (SD: 0.09), respectively, in Jam, while, on the liability scale, they are 0.06 (SD: 0.04) and 0.05 (0.04) (test statistic = 17.71, *P*_permutation_ = 7.38E-06 for *BOP1*, and test statistic = 16.83, *P*_permutation_ = 1.08E-05 for *HSF1*). The *BOP1* and *HSF1* results coincide with the gene enrichment findings above in Jam. *GALNT13* harbouring the replicated rs10196189 yielded a heritability estimate of 0.19 (SD: 0.08) on the observed scale and 0.11 (SD: 0.04) on the liability scale in Jam (test statistic = 9.14, *P*_permutation_ = 7.81E-04), in contrast to the negligible heritability estimated in A-A (test statistic = 0.02, *P*_permutation_ = 0.32) and Jpn (test statistic = 0, *P*_permutation_ = 0.68). It should be noted that these observations in Jam were not statistically significant, after accounting for the number of genes analyzed.

### Functional genomic annotations and gene re-prioritization

In the final phase, we performed tissue-specific functional network analysis in GIANT^29^, functional module analysis^30^, Sei analysis^31^ of genomic variant effects on regulatory features and additional disease-gene association analysis in NetWAS^29^ based on the SNP- and gene-based results; implemented in HumanBase (https://humanbase.io/; Methods). We focused on the nine genes and their sentinel variants, where appropriate, including *GALNT13*, *STXBP2*, *BOP1*, *HSF1*, *GRM7*, *MPRIP*, *CERS4*, *ZFYVE28* and *ADAMTS18*, for GIANT and Sei analyses. One hundred ninety-nine genes enriched in the interferon-gamma response gene-set were taken forward for the functional module detection. The gene-based results were also fed into NetWAS for gene reprioritization for the additional disease-gene association analysis.

### Functional networks (GIANT)

Among *GALNT13*, *STXBP2*, *BOP1*, *HSF1*, *GRM7*, *MPRIP*, *ZFYVE28* and *ADAMTS18* analyzed in GIANT, tissue-/cell-type specific expression networks revealed that *GALNT13, GRM7*, *MPRIP, ZFYVE28,* and *ADAMTS18* are expressed almost universally across the nervous system (Figure 2a). The highest predicted confidence score is observed in *ZFYVE28* for its expression in caudate nucleus (score: 0.87, Figure 2a). *STXBP2* is expressed universally across the hematopoietic system (score: 0.15 – 0.87), while its expression is experimentally validated in blood platelet (score: 1, Figure 2b). Intriguingly, the expression of *MPRIP* is observed across multiple tissue systems, including the nervous, muscular, endocrine, cardiovascular, skeletal, and embryonic systems, being experimentally validated in smooth muscle (Figure 2c).

**Figure 2.**
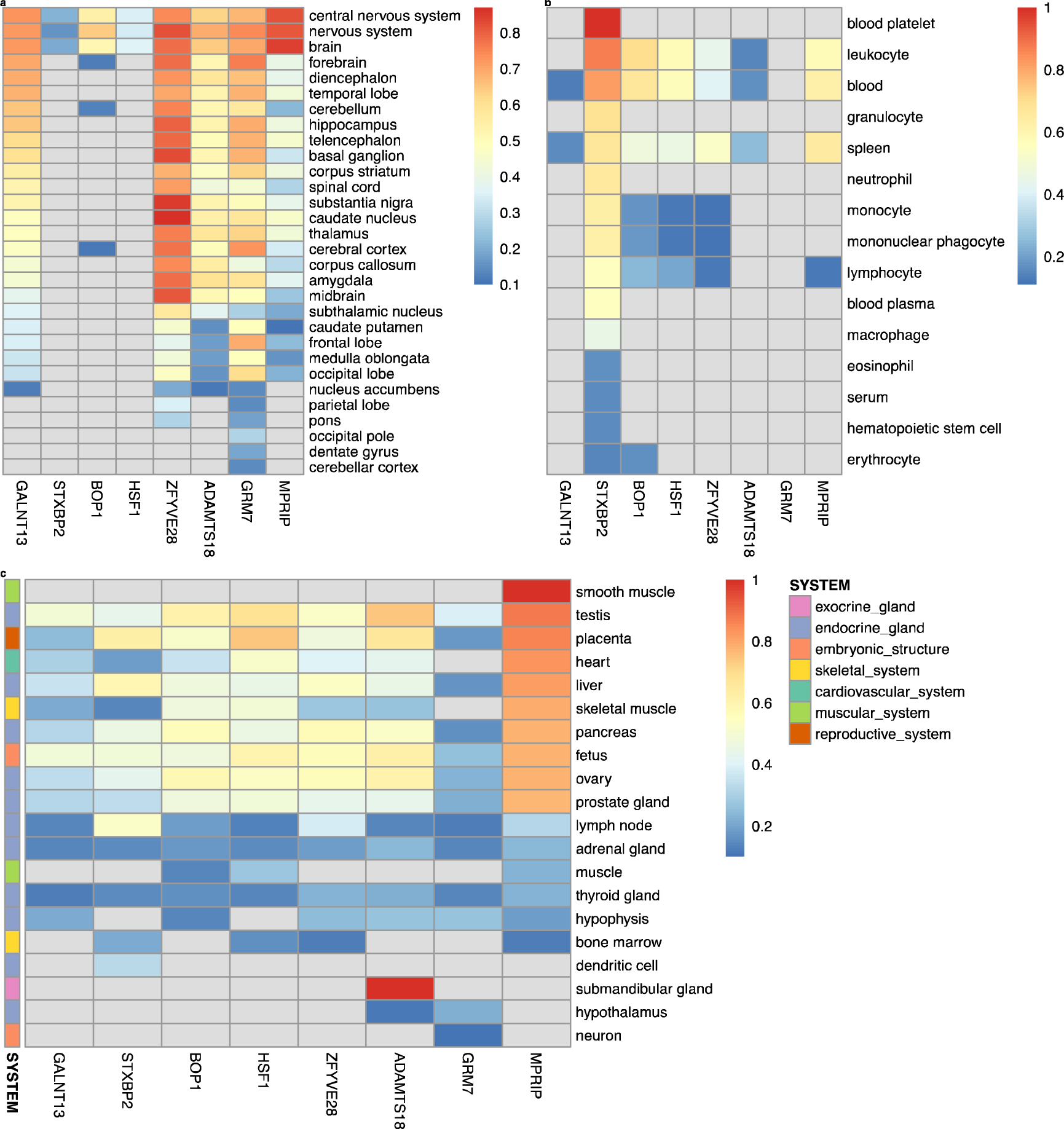
Tissue-/cell-type specific expression of *GALNT13*, *STXBP2*, *BOP1*, *HSF1*, *ZFYVE28*, *ADAMTS18*, *GRM7* and *MPRIP* in the nervous system (a), hematopoietic system (b), as well as other multiple systems (c), following the functional networks analysis in GIANT.

### Polypeptide N-acetylgalactosaminyltransferase 13 (*GALNT13*)

Tissue-specific gene expression in the central nervous system was noted for *GALNT13* (highest predicted score: 0.73) relative to other nervous tissue types (Figure 2a). In this network, 14 genes are functionally similar to *GALNT13* based on their interaction score (> 0.15), including *GNAO1*, *ASIC1*, *GRIK2*, *MED12L*, *SORCS1*, *GRIK3*, *FZD4*, *SEZ6L*, *MYO7A*, *SHANK2*, *KMT2A*, *KANSL1*, *DSCAM*, and *PIEZO2* (Supplementary Fig. S4). Seven of them are associated with the following biological processes represented in the network: the nervous system process (*ASIC1*, *GRIK2*, *MYO7A* and *SHANK2*), sodium ion transmembrane transport (*ASIC1*, *GRIK2*, and *MYO7A*), histone H4-K16 acetylation (*KMT2A* and *KANSL1*), ionotropic glutamate receptor signalling pathway (*GRIK2* and *GRIK3*), and monovalent inorganic cation transport (*ASIC1*, *GRIK2* and *GRIK3*) (*P* < 3.70E-02). Most gene pairs of *GALNT13* in the network also show evidence for their role in locus ceruleus (a nucleus in the pons of the brain stem, which activates the sympathetic nervous system) (interaction score: 0.36 – 0.91; Supplementary Data S1).

For biological process-specific gene interactions, we observed *GALNT13*–*TF* (transferrin) gene pair associated with forebrain development (highest interaction score: 0.89), followed by *GALNT13*–*FUT6*, *GALNT13*–*SIX3*, *GALNT13*–*GCNT3*, *GALNT13*–*BTG4*, and *GALNT13*–*RTL3* (interaction score: 0.51 – 0.64; Supplementary Fig. S4). In addition, dorsal/ventral pattern formation, regionalization and pattern specification process are enriched in forebrain development (*P* < 3.80E-02), marked by the network genes *SIX3* and *SMAD6*.

### Syntaxin binding protein 2 (*STXBP2*)

The highest predicted score of 0.87 for *STXBP2* expression was found in leukocyte (Figure 2b). The top and functionally similar genes to *STXBP2* consist of *CAPG*, *PTPN6*, *S100A11*, *EHBP1L1*, *IFI30*, *ARPC4*, *PTPN18*, *MVP*, *ACTN4*, *CYBA*, *TYMP*, *SBNO2*, *PFN1*, and *IKBKG* (interaction score > 0.46; Supplementary Fig. S4). Gene pairs of *STXBP2* in the network show evidence supporting their functions in osteoblasts, granulocytes (basophils), monocytes, dendritic cells, natural killer cells, locus ceruleus and dentate gyrus (interaction score: 0.51 – 0.93; Supplementary Data S1). Gene interactions specific to the biological process of leukocyte migration are characterized by *STXBP2*–*ARHGEF1*, *STXBP2*–*CNOT3*, *STXBP2*–*CAPG*, *STXBP2*–*SBNO2*, *STXBP2*–*GRN*, *STXBP2*–*CLDN4, STXBP2*–*MVP, STXBP2*–*STX4* and *STXBP2*–*POLD4* (interaction score ≥ 0.95; Supplementary Fig. S4). Particularly, *STXBP2*, *SBNO2* and *STX4* are associated with several biological processes enriched in leukocyte migration, such as myeloid cell and leukocyte activation involved in immune response (*P* = 1.00E-03 and *P* = 4.00E-03, respectively), and granulocyte activation (*P* = 1.40E-02).

### BOP1 ribosomal biogenesis factor (*BOP1*) and heat shock transcription factor 1 (*HSF1*)

*BOP1* shows the highest expression in leukocyte, followed by the blood and the nervous system (predicted score: 0.70, 0.67, and 0.64, respectively) as well as in endocrine gland, such as testis, ovary, and pancreas (predicted score: 0.61, 0.58, and 0.56, respectively) (Figure 2). *HSF1* is most expressed in the reproductive system (placenta, 0.74), endocrine gland (testis, 0.68) and embryonic structure (fetus, 0.60), followed by leukocyte (0.58), blood (0.57), heart (0.52), skeletal muscle (0.49), brain (0.36), and other endocrine glands (ovary, 0.54; prostate gland, 0.48; pancreas, 0.46; liver, 0.45) (Figure 2). *BOP1* and *HSF1* are physically close and located on chromosome 8. The interaction confidence between them is the strongest in peripheral nervous system (0.97). Evidence for this prediction probability comes from gene co-expression (62%), TF binding prediction (26%) and GSEA perturbations datasets (12%). Other genes in the network that are functionally similar to *BOP1* and *HSF1* are comprised of *FGFR3*, *TRIP6*, *PTBP1*, *FGFR1*, *CCNF*, *UPF1*, *SLC52A2*, *SCRIB*, *GAL*, *C20orf27*, *NUP188*, *EIF3B* and *SMARCA4* (interaction score: 0.88 – 0.96) (Supplementary Fig. S4). Among them, 5 genes are associated with viral gene expression (*PTBP1*, *EIF3B* and *SMARCA4*), IRES-dependent viral translation initiation (*PTBP1* and *EIF3B*), synostosis (*FGFR3* and *FGFR1*), and viral translation (*PTBP1* and *EIF3B*) enriched in the peripheral nervous system (*P* ≤ 2.80E-02). Gene pairs of *BOP1* or *HSF1* in the network are also predicted to function in hepatocytes, corpus luteum, trophoblast, cardiac muscle, adrenal cortex, and uterine cervix (interaction score: 0.56 – 1.00; Supplementary Data S1).

In addition, *BOP1* and *HSF1* are specifically involved in the ubiquitin-dependent protein catabolic process (Supplementary Fig. S4). Top three genes connected to *BOP1* and *HSF1*, i.e., *ARHGDIA*, *EIF4G1* and *ATP5F1D* (interaction score > 0.90), show experimentally validated associations with glutamate receptor signalling, ribonucleoprotein complex assembly, translation initiation, cellular respiration and rRNA processing. *BOP1* and *DHX30* are also associated with ribosomal large subunit assembly (*P* = 1.50E-02) and ribosomal biogenesis (*P* = 2.4E-02), which are enriched in the ubiquitin-dependent protein catabolic process.

### Glutamate metabotropic receptor 7 (*GRM7*)

The *GRM7* gene is universally expressed across the nervous tissue system, ranging from the lowest expression in parietal lobe, nucleus accumbens and cerebellar cortex (predicted score: 0.14) to the highest expression in forebrain (0.76) (Figure 2a). Functionally similar genes to *GRM7* in the forebrain include *ATP2B2*, *MYT1L*, *GRIK3*, *LRP6*, *EDA*, *PKNOX2*, *CACNA1I*, *CATSPER2*, *NELL1*, *AFF2*, *GABRA4*, *DOCK1*, *PAPPA2*, and *RALYL* (interaction score: 0.28 – 0.41) (Supplementary Fig. S4). Among them, *GRM7* and *GRIK3* are present in several biological processes enriched in the forebrain network and involved in neurotransmission, such as adenylate cyclase-inhibiting G-protein coupled glutamate receptor signalling pathway (*P* = 7.5E-04) and G-protein coupled receptor signalling pathway, coupled to cyclic nucleotide second messenger (*P* = 4.40E-02). In addition, *GRM7* and *ATP2B2* are found in sensory perception of sound and mechanical stimulus (*P* = 2.20E-02). The majority of gene pairs of *GRM7* in the network are also being supported for their role in locus ceruleus (interaction score: 0.68 – 1; Supplementary Data S1).

For biological process-specific gene interactions in the glutamate receptor signalling pathway, *GRM7*, *GRM1*, *GRIK3*, *GRIK1*, *GRIA1*, *GRIA2*, *GRIN1*, and *GRMB* are over-represented in this network (*P* = 1.00E-12). In addition, among the network genes, *GRIK3*, *GRIK1*, *GRIA1*, *GRIA2*, and *GRIN*1 associate with ionotropic glutamate receptor signalling pathway (*P* = 5.20E-09), *GRM1*, *GRM7*, *GRM8*, and *GRIK3* with G-protein coupled glutamate receptor signalling pathway (*P* = 6.30E-09), and *GRM7*, *GRM8* and *GRIK3* with adenylate cyclase-inhibiting G-protein coupled glutamate receptor signalling pathway (*P* = 2.60E-07) (Supplementary Fig. S4).

### Myosin phosphatase Rho interacting protein (*MPRIP*)

*MPRIP* is expressed across multiple tissue systems; notably, in smooth muscle, endocrine gland (e.g., testis, liver, pancreas, ovary, and prostate gland), placenta, skeletal muscle and the nervous system (Figure 2). In these tissue networks, *MPRIP* and *YWHAB* strongly interact in smooth muscle (interaction score: 0.71), testis and placenta (0.82), skeletal muscle (0.73), and brain (0.61). The gene-pair is also predicted to function in spermatogonium (0.96), parietal lobe (0.95), pons (0.93), eosinophil (0.92), spermatid (0.92) and T-lymphocyte (0.89).

In addition, the pair plays their part in a number of specific biological processes and pathways (interaction score ≥ 0.9), such as, in the descending order of the interaction score, translational initiation, gene silencing by RNA, RNA splicing, cell adhesion mediated by integrin, cellular senescence, actin cytoskeleton reorganisation, acute inflammatory response, T cell proliferation, cell morphogenesis, erythrocyte differentiation, cardiac muscle contraction, extracellular matrix organisation, telomere maintenance, Ras protein signal transduction, neuron projection morphogenesis, response to hypoxia, chemical synaptic transmission, necrotic cell death, angiogenesis, actin filament bundle assembly, T-cell differentiation, T-cell mediated immunity, and response to virus.

On tissue-specific level and for genes functionally similar to *MPRIP*, *YWHAB* and *FLNA* are enriched in cytoplasmic sequestering of protein (smooth muscle, testis and placenta, *P* ≤ 3.00E-02), *PAFAH1B1* and *FLNA* in congenital nervous system abnormality (testis, *P* = 3.10E-02), *ACTN1*, *MACF1*, *SMAD3* and *PPM1F* in cell-cell junction organisation (placenta, *P* = 1.60E-03), *ACTN1*, *MACF1*, and *PPM1F* in cell-substrate adherens junction assembly and cell-matrix adhesion (placenta, *P* = 1.60E-03 and 7.50E-03, respectively), *ACTN1*, *PPM1F*, and *FLNA* in actin filament organisation and actin cytoskeleton organisation (placenta, *P* = 1.60E-02 and 3.90E-02, respectively), *PPM1F* and *ITGB1BP1* in cell-substrate adherens junction assembly (brain, *P* = 2.90E-02), *ITGB1BP1* and *TAX1BP3* in negative regulation of cellular protein localization (brain, *P* = 3.90E-02), *LAMC1*, *RHOA* and *MACF1* in cell junction assembly and cell junction organisation (skeletal muscle, *P* = 3.40E-02), and *RHOA*, *MACF1* and *YWHAG* in regulation of neuron differentiation, regulation of neurogenesis and regulation of nervous system development (skeletal muscle, *P* ≤ 4.60E-02).

### GALNT13, GRM7 and MPRIP

These three genes originate from the Jam (*GALNT13*), A-A (*GRM7*) and Jpn (*MPRIP*) cohorts, crossing the suggestive threshold of *P* = 5E-05 in the discovery GWAS. They are also almost universally expressed across the entire nervous tissue system (Figure 2a). We therefore focused on the three genes solely examining their interactions in the nervous system. They are most strongly characterized in the glutamate receptor signalling pathway: minimum interaction score was set to 0.20 for filtering the gene network and only the top 10 genes in the network are considered here given their uniqueness to the query genes, including *PCDHGC5*, *OR4D2*, *TAS2R50*, *KNG1*, *ANKRD31*, *KRT82*, *TEX26*, *FOXR2*, *MGAT4C* and *EXOC3L4* (Supplementary Fig. S4).

*GALNT13* and *GRM7* have an interaction score of 0.33 in the glutamate receptor signalling pathway (interaction evidence: 79% co-expression, 20% GSEA perturbations, and 1% TF binding), while they most strongly interact in chemical synaptic transmission (0.80). The majority of gene pairs of *GALNT13* or *GRM7* are co-expressed in glutamate receptor signalling pathway, whereas there are also functional evidence supporting their specific roles in chemical synaptic transmission, detection of chemical stimulus, sodium ion transport, potassium ion transport, hormone secretion, cardiac ventricle development, cardiac muscle contraction, and cAMP biosynthetic process (interaction score: 0.29 – 1.00; Supplementary Data S1). Direct interactions for *MPRIP* were observed for *MPRIP*–*ANKRD31*, *MPRIP*– *EXOC3L4* and *MPRIP*–*PCDHGC5* in chemical synaptic transmission (interaction score: 0.26), cartilage development (0.47) and cardiac ventricle development (0.58), respectively; in addition to their functional relationship to glutamate receptor signalling pathway (interaction score: 0.27, 0.50 and 0.69, respectively) (Supplementary Data S1).

### Zinc finger FYVE-type containing 28 (*ZFYVE28*)

Tissue-specific expression of *ZFYVE28* is observed across most parts of the nervous system, being the highest in caudate nucleus (predicted score: 0.87), followed by substantia nigra (0.85) and basal ganglion (0.83) (Figure 2a). In caudate nucleus, *CHD9*, *PRNP*, *PSMB7*, *SUPT7L*, *USP46*, *ACOT11* and *TTC17* interact with *ZFYVE28* (interaction score: 0.16 – 0.20, Supplementary Fig. S4), and these interactions are also captured in amygdala (0.17 – 0.20). In addition, interactions between *PSMB7*, *USP46*, *ACOT11* and *ZFYVE28* are observed in substantia nigra (0.18 – 0.19), and between *CHD9*, *PRNP*, *TTC17* and *ZFYVE28* in basal ganglion (0.18 – 0.21). Most of these gene pairs show stronger interactions in locus ceruleus (0.29 – 0.72; Supplementary Data S1).

### ADAM metallopeptidase with thrombospondin type 1 motif 18 (*ADAMTS18*)

*ADAMTS18* is expressed in submandibular gland (Figure 2c), followed by its ubiquitous expression across the nervous tissue system with the highest expression being in the nervous system (predicted score: 0.69, Figure 2a). Top genes functionally similar to *ADAMTS18* in the nervous system include *INHBB*, *SLC9A2*, *ITGA11*, *BCL2*, *SMOC2*, *CSMD1*, *BAIAP2*, *PDGFRA*, *JPH3*, *ACTL6B*, *WNT1*, *TBC1D26*, *CHRD* and *GDF6* (interaction score: 0.16 – 0.19) (Supplementary Fig. S4). Among them, *INHBB*, *PDGFRA*, *WNT1* and *ADAMTS18* are enriched in response to wounding (*P* = 2.10E-03); *INHBB*, *WNT1*, *CHRD* and *GDF6* in transmembrane receptor protein serine/threonine kinase signalling (*P* = 2.10E-03); *WNT1*, *CHRD* and *GDF6* in BMP signalling pathway and cellular response to BMP stimulus (*P* = 2.10E-03), *ADAMTS18* and *PDGFRA* in negative regulation of platelet activation (*P* = 2.10E-03) and aggregation (*P* = 6.10E-03), *PDGFRA*, *WNT1*, *CHRD* and *GDF6* in cellular response to growth factor stimulus (*P* = 6.10E-03), *PDGFRA*, *WNT1*, *ADAMTS18*, and *BAIAP2* in cell-cell adhesion (*P* = 6.10E-03), *PDGFRA* and *WNT1* in positive regulation of fibroblast proliferation (*P* = 8.60E-03), *PDGFRA*, *BCL2*, and *ADAMTS18* in regulation of cell activation (*P* = 1.30E-02) and *WNT1* and *GDF6* in skeletal system development (*P* = 4.70E-02). Four gene pairs, *ACTL6B*–*ADAMTS18*, *JPH3*–*ADAMTS18*, *TBC1D26*–*ADAMTS18* and *CSMD1*–*ADAMTS18* also closely interact in locus ceruleus (interaction score: 0.24, 0.51, 0.63 and 0.88, respectively; Supplementary Data S1).

### Blood-specific functional modules

The competitive gene-set enrichment analysis above revealed that 199 genes in the gene-set of interferon-gamma response are significantly enriched in the meta-analysis results. We now focus on identifying blood-specific functional modules underlying this gene set. In this analysis, 109 genes (out of 197 genes, excluding *WARS1* and *MARCHF1* not matched in the humanbase data collections) were assigned to a specific module, i.e., M1-M7 (Figure 3). The top three representative biological processes, annotated to Gene Ontology terms, in each module, consist of response to virus, defense response to virus, and defense response to other organism (M1, 38 genes; Q < 1.00E-4); viral life cycle, regulation of viral genome replication, and regulation of viral life cycle (M2, 40 genes; Q < 1.00E-4); regulation of endothelial cell apoptotic process, endothelial cell apoptotic process and regulation of epithelial cell apoptotic process (M3, 8 genes; Q < 1.00E-4); regulation of response to interferon-gamma, regulation of interferon-gamma-mediated signalling pathway, and interferon-gamma-mediated signalling pathway (M4, 4 genes; Q < 1.00E-4); leukocyte migration, natural killer cell chemotaxis, and regulation of natural killer cell chemotaxis (M5, 15 genes; Q = 1.00E-4); positive regulation of endopeptidase activity, positive regulation of peptidase activity, and positive regulation of proteolysis (M6, 2 genes; Q ≤ 8.00E-4); and muscle cell proliferation, regulation of inflammatory response and inflammatory response (M7, 3 genes; Q ≤ 8.00E-4). Among the 109 genes, 8, 18, 3, 1, and 6 genes are uncharacterized in M1-M5, respectively (totalling, 36 genes); in other words, between approximately 21% and 43% of genes within each module (M1-M5) lack any annotation to specific Gene Ontology terms, indicative of their novel roles in the specific functional modules (Figure 3). Notably, *IFI35*, *PSME2*, *HLA-A* and *UBE2L6* are the top-ranked uncharacterized genes in M2, *IL2RB* and *FGL2* in M5, and *IFI44* in M1 (Figure 3). In M2, the uncharacterized pairs of *PSME2*–*PSME1* and *HLA-A*– *HLA-B*, display strong edge weights of 0.997 and 0.987, respectively. In addition, *IFI35* associates with 11 other genes in the local network (including 6 other uncharacterized genes: *IFI30*, *LGALS3BP, LY6E*, *NMI*, *PARP12* and *UBE2L6*; Figure 3), and the overall edge weight ranges from 0.981 (*IFI35*–*ISG15*) to 0.816 (*IFI35*–*MVP*) (Supplementary Data S2).

**Figure 3.**
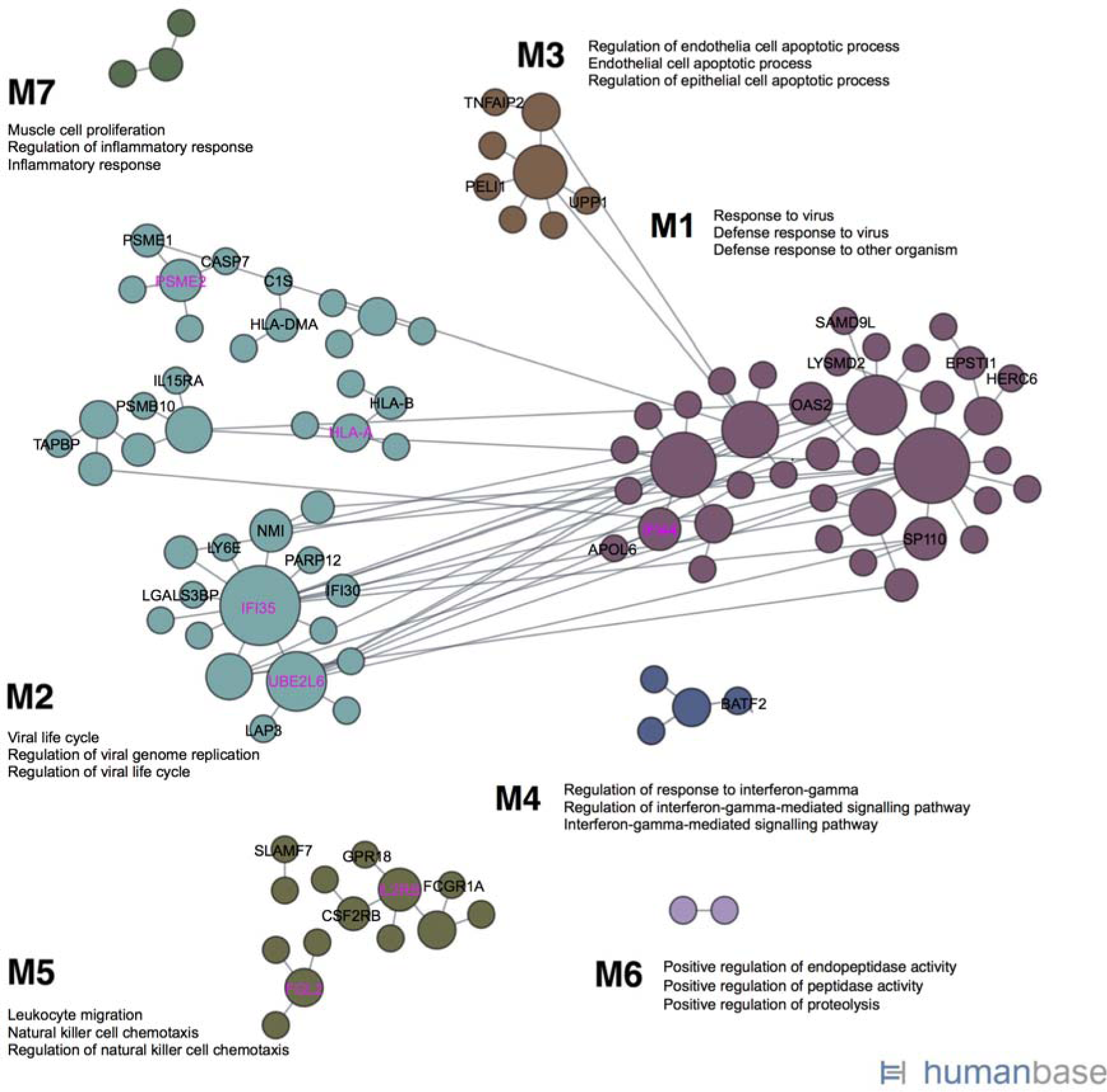
Cellular responses to interferon-gamma unveiled in blood-specific functional modules, linking individual genes to specific biological processes. Top three enriched GO terms are annotated for each module, and novel, previously uncharacterized genes are labelled in M1-M7 (among which, the highest ranked genes are marked in magenta based on the number of edges they associate with). The figure is adapted from the output of the modules analysis implemented in humanbase. Each node represents a gene and the association between any gene pair is denoted by an edge.

### Genomic variant effect analysis (Sei analysis)

The variant effects of six sentinel SNPs representing independent regional signals near or in *GLANT13*, *STXBP2*, *BOP1*, *GRM7*, *MPRIP*, *CERS4* on tissue-/cell-type-specific regulatory activities were quantified in Sei (Table 4). The six SNPs are resulted from the discovery GWAS crossing the suggestive cut-off of *P* = 5E-05 (rs2303115, *STXBP*, Jam; or replaced by the stronger Sanger imputation signals in almost perfect LD with them: rs4977199, *BOP1*, *P* = 7.73E-07, Jam; and rs2875287, *GRM7*, *P* = 3.79E-07, A-A) or represent the top associations following imputation and/or meta-analysis (rs117143557, *MPRIP*, Jpn, *P*_GC-corrected_ = 7.63E-08; rs2927712, *CERS4* across Jam and A-A, *P*_GC-corrected_ = 2.08E-07; and rs113303758, *GALNT13* across Jam, A-A, and Jpn, *P*_GC-corrected_ = 3.16E-07).

**Table 4.**
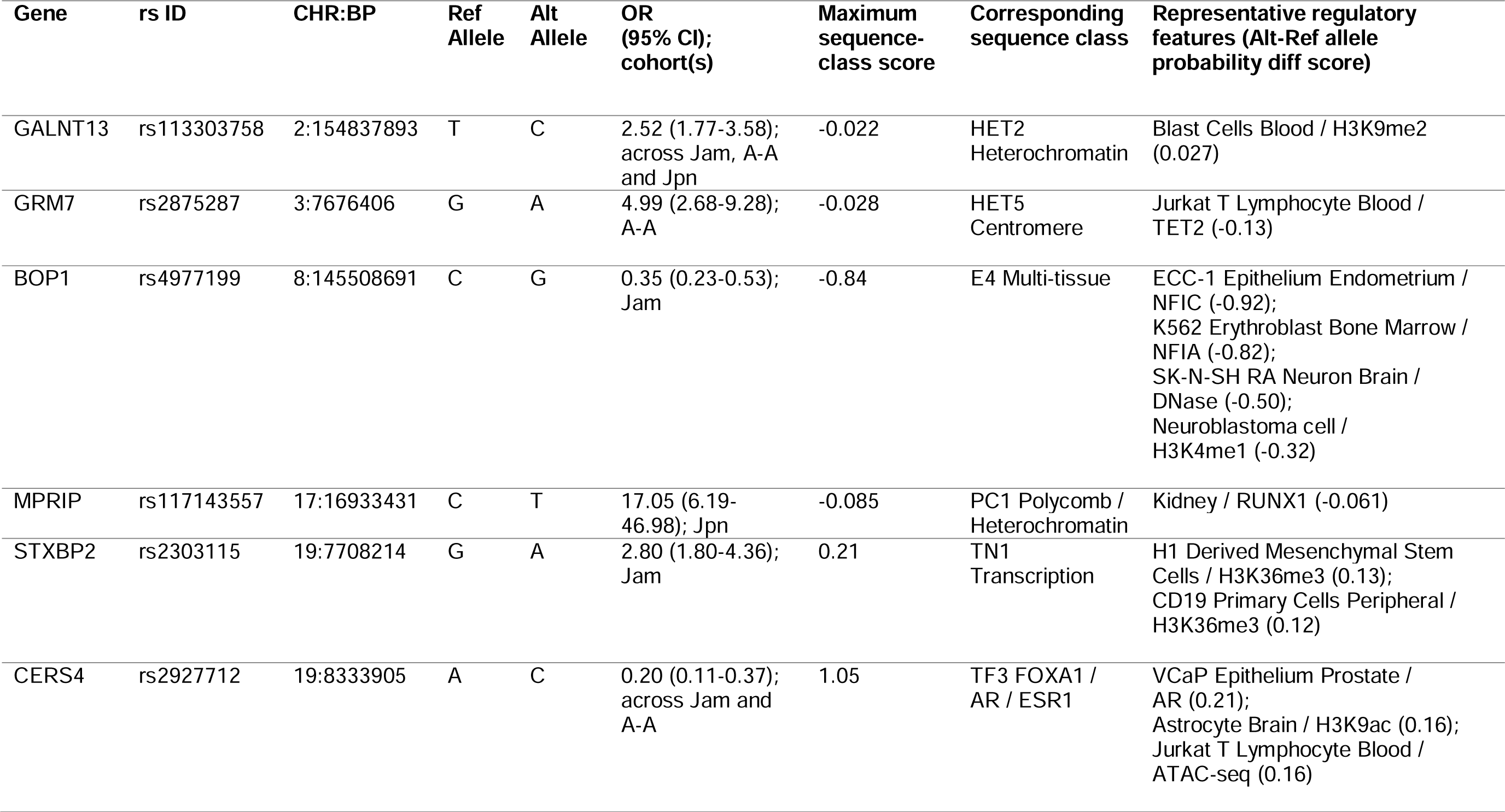
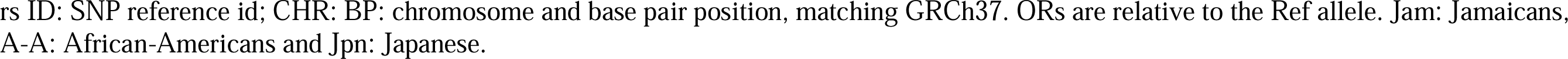
Tissue-/cell-type-specific regulatory activities associated with sentinel SNPs resided within or next to genes derived from the SNP- and gene-based analysis in Sei (predictions made for 40 sequence classes and 21,907 regulatory features).

#### Sequence class-level variant effects

The C-allele of rs2927712 (intergenic variant, nearest gene *CERS4*) increases the activities of the transcription factor (TF) sequence classes TF3 FOXA1/AR/ESR1 (maximum sequence class score: 1.05), TF5 AR (0.97), and the enhancer (E) class E4 Multi-tissue (0.72); the C-allele decreases the activity of E5 B-cell-like (−0.50). The A-allele of rs2303115 (intron variant in *STXBP2*) increases the activities of transcription (TN) sequence classes TN1 (0.21), TN2 (0.18) and TN4 (0.17). The G-allele of rs4977199 (intron variant in *BOP1*) mitigates the activities of several enhancer classes—notably, E4 Multi-tissue (−0.84), E3 Brain/Melanocyte (−0.64), and E1 Stem cell (−0.62), P promotor (−0.56), and TF3 FOXA1/AR/ESR1 (−0.50)— but increases the transcription of TN1 (0.70), TN4 (0.69) and TN2 (0.45).

The C-allele of rs113303758 (intron variant in *GALNT13*) overall decreases the activities of the heterochromatin (HET) sequence classes HET2 (−0.022) and HET1 (−0.020). The A-allele of rs2875287 (intron variant in *GRM7*) decreases the activities of HET5 Centromere (−0.028) and HET6 Centromere (−0.018), alongside the increased activity of HET4 heterochromatin (0.019) and HET3 heterochromatin (0.015). The T-allele of rs117143557 (intergenic variant, nearest gene *MPRIP*) decreases all sequence classes activity, particularly the activity of Polycomb (PC) sequence class PC1 Polycomb/Heterochromatin (−0.085).

#### Regulatory feature scores for 21,907 chromatin profiles

On the level of regulatory features, the C-allele of rs2927712 shows higher probabilities of increasing the androgen receptor activity in VCaP Epithelium Prostate (maximum alt-ref allele diffs score: 0.21), the H3K9ac-marked transcription activity in Astrocyte Brain (0.16) and the chromatin accessibility in Jurkat T Lymphocyte Blood (0.16). The A-allele of rs2303115 displays a higher probability of increasing H3K36me3-marked transcription activity in Fetal Stomach (0.18), Fetal Intestine Small (0.15), Fetal Muscle Leg (0.14), H1 Derived Mesenchymal Stem Cells (0.13), CD19 Primary Cells Peripheral (0.12), Monocytes-CD14+ RO01746 (0.11). The G-allele of rs4977199 decreases the activities of *NFIC* in ECC-1 Epithelium Endometrium (−0.92), *NFIA* in K562 Erythroblast Bone Marrow (−0.82), DNase in SK-N-SH RA Neuron Brain (−0.50) and heart (−0.35), and H3K4me1-marked enhancer activity in Neuroblastoma cell (−0.32) and Brain Cingulate Gyrus (−0.16), while increases the activities of *CNOT3* in HCT-116 Colorectal cancer cell line (0.20), *CBFB* in ME-1 Leukaemia cell (0.13), H3K36me3-marked transcription in Left Ventricle (0.12) and Adipose Nuclei (0.12) and H3K27me3-marked repression in Astrocyte (0.12).

The C-allele of rs113303758 is linked to H3K9me2-marked epigenetic repression in Blast Cells Blood (0.027) and K562 Erythroblast Bone Marrow (0.019). The A-allele of rs2875287 associates with the reduced activities of *TET2* in Jurkat T Lymphocyte Blood (−0.13) and H3K27me3-marked repression in PAR Astrocyte (−0.091) but increases the activity of H3K27me3 in Jurkat T Lymphocyte Blood (0.060). The T-allele of rs117143557 has a variety of functional roles, for example, linking to H3K4me3-marked promoter activity in IMR90 Fibroblast Lung (0.060), H3K27ac-marked enhancer activity in Monocyte Blood (0.055), H3K27me3-marked repression in Proerythroblast Bone Marrow (0.041), and H3K18ac-marked DNA demethylation^32^ in H1 Derived Neuronal Progenitor Cultured Cells (0.038) while decreasing the activities of the transcription factor RUNX1 in kidney (−0.061) and the repressive H3K27me3 marks in PAR Astrocyte (−0.051).

### GWAS reprioritization for disease-gene associations (NetWAS)

The NetWAS analysis reprioritized 14,788 (Jam), 15,472 (A-A), 1,846 (Jpn) and 13,301 genes (meta-analysis)—characterized by positive NetWAS scores, thereby likely being nominally significant—in the relevant tissue networks. To evaluate these reprioritized gene lists, we then submitted them to Enrichr^33–35^ for functional annotations focusing on gene enrichment in “Diseases/Drugs” and “Cell Types” gene-set libraries.

Using the central nervous system network, the 14,788 genes reprioritized from the Jam cohort are significantly enriched, such as, for top neuron-specific and neuron-enriched genes in humans and mice (OR = 1.96, Q = 7.64E-13, neuron-specific and OR = 1.82, Q = 1.55E-10, neuron-enriched), genes changed in striatum and cortex of HdhQ175 mice (OR = 1.45, Q = 4.46E-11 striatum and OR = 1.52, Q = 1.65E-08 cortex), H3K27me3-enriched genes in MSNs of adult mice (OR = 1.43, Q = 5.29E-08) and differentially expressed genes in hippocampus of Foxp1 knock-out versus wild-type mice (OR = 1.40, Q = 1.29E-07) in the HDSigDB Human 2021 gene sets. A full list of enrichment terms, overlapping genes, raw P-value, Q-value, OR and Enrichr combined score for the HDSigDB Human 2021 terms is available in Supplementary Data S3. Genes from this list are also highly expressed across a range of brain regions and the spinal cord in the ARCHS4 Tissues gene sets (Supplementary Data S4), in glutamatergic neuron in the CellMarker Augmented 2021 gene-set library (OR = 6.66, Q = 5.00E-05) and in the adult frontal cortex gene-set from ProteomicsDB (OR = 1.97, Q = 6.00E-05).

In the forebrain network, the 15,472 genes reprioritized from the A-A cohort are enriched for developmental transcription factor genes bound by Suz12 (OR = 1.89, Q = 4.10E-02), which regulate early developmental processes, for example, neurogenesis, haematopoiesis and cell-fate specification, in HDSigDB Human 2021 (Supplementary Data S5), and for genes expressed in GABAergic neurons from the single-cell RNA-seq database PanglaoDB (OR = 2.04, Q = 2.00E-02), but more so in macrophages (OR = 2.44, Q = 1.60E-03), dendritic cells (OR = 2.37, Q = 1.70E-03), hematopoietic stem cells (OR = 2.09, Q = 7.60E-03) and the resident macrophage cells, microglia, in the brain and spinal cord (OR = 2.15, Q = 9.50E-03) (Supplementary Data S6).

Using the brain network, 1,846 genes reprioritized from the Jpn cohort are enriched for NP2 neural progenitor cell-enriched genes (OR = 1.64, Q = 4.10E-17) and genes down-regulated in Sdt/Npy-expressing interneurons of HD patients versus controls (OR = 1.62, Q = 2.90E-16) in HDSigDB Human 2021 (Supplementary Data S7). This gene list also significantly overlaps with genes expressed in prefrontal cortex (OR = 1.76, Q = 1.10E-03), CD8+ T cells (OR=1.53, Q=0.020) and cerebellum peduncles (OR = 2.21, Q = 2.00E-02) in the Human Gene Atlas gene-sets (Supplementary Data S8), in fibrous astrocytes (Human Astro L1-6 FGFR3 AQP1 down, OR = 2.08, Q = 1.99E-08) from the Allen Brain Atlas 10x scRNA 2021 library and in retrosplenial area, lateral agranular part, and layer 6b neurons (OR = 2.03, Q = 3.40E-02) from Allen Brain Atlas (Supplementary Data S9).

Using the tissue network for caudate nucleus, 13,301 genes reprioritized given the meta-analysis are also enriched for top neuron-enriched and neuron-specific genes in humans and mice (OR = 1.52, Q = 8.75E-06, neuron-enriched and OR = 1.36, Q = 4.60E-03 neuron-specific), genes changed in brain versus spinal cord derived OPCs (OR = 1.41, Q = 2.60E-03), genes up-regulated in hippocampus of Foxp1 knock-out versus wild-type mice (OR = 1.29, Q = 2.70E-04) and genes up-regulated in Pvalb/Th-expressing interneurons of zQ175DN versus WT (OR = 2.22, Q = 8.40E-03) in the HDSigDB Human 2021 gene sets (Supplementary Data S10). This gene list also entails genes highly expressed across the whole brain, superior frontal gyrus, prefrontal cortex, motor neuron, cerebral cortex and the spinal cord in the ARCHS4 Tissues gene sets (Supplementary Data S11), in layer 1 of AOV cortex, layer 1 of FCx, mantle zone of AOV, superficial stratum of AOV, Forel’s field, and nucleus of the posterior commissure in the Allen Brain Atlas gene-sets (Supplementary Data S12) and in human motor cortex in HuBMAP ASCTplusB augmented 2022 gene-sets (Supplementary Data S13).

Additionally, focusing on the four genes including *UQCRFS1* (Jam), *PTPN6* (A-A), *RALY* (Jpn), and *ZMYM4* (meta-analysis) with the highest positive NetWAS score from each cohort (Supplementary Data S14), a knowledge search of these genes in Enrichr gene-set libraries revealed that knockdown of *UQCRFS1* results in extended life span in female mice (MP:0001661) and in contrast, results in premature death in male mice (MP:0002083) in MGI Mammalian Phenotype Level 4 2021; and *UQCRFS1* is down-regulated in human (GSE53890), rat (GDS3939) and mouse (GSE20411) brain (frontal cortex) during aging in Aging Perturbations from GEO libraries. Knockdown of *PTPN6* results in abnormal astrocyte morphology (MP:0002182), abnormal forebrain morphology (MP:0000783), abnormal nervous system morphology (MP:0003632), decreased brain size (MP:0000774), abnormal hippocampus development (MP:0000808), and decreased neuron number (MP:0008948) among others, such as decreased apoptosis (MP:0006043), decreased bone mass (MP:0004016), and increased erythroid progenitor cell number (MP:0003135), in MGI Mammalian Phenotype Level 4 2021. Knockdown of *RALY* results in abnormal lung morphology (MP:0001175) and short tibia (MP:0002764) in MGI Mammalian Phenotype Level 4 2021. *RALY* is up-regulated in Claustrum cells and in the primary visual area, layer 5 cells in Allen Brain Atlas gene-sets. Knockdown of *ZMYM4* results in decreased body length (MP:0001258), decreased lean body mass (MP:0003961), and decreased mean platelet volume (MP:0008935) in MGI Mammalian Phenotype Level 4 2021. In addition, *ZMYM4* is down-regulated in Alzheimer’s Disease (GSE1297), Parkinson’s Disease (GSE7621), chronic obstructive pulmonary disease (GSE3320) and large granular lymphocytic leukaemia (GSE10631) in Disease Signatures from GEO 2014.

## Discussion

Here, we conducted a cross-ancestry, multiphase GWAS in three cohorts of world-class sprint athletes and their matched controls. We carried out both the SNP- and gene-based analyses, followed by machine-learning based functional characterization and reprioritization of disease-gene associations. Specifically, the imputation pipeline significantly boosted the strength of GWAS discoveries in the relatively small number of samples available to this study (Supplementary Fig. S2), resulting in stronger and novel signals (Supplementary Figures S5-S9 for *GALNT13*, *BOP1*, *HSF1*, *GRM7*, *MPRIP* and *STXBP2*). Independent replication for rs10196189 (*GALNT13*) and meta-analysis further boosted the power of the study in capturing genetic associations for *GALNT13, ZFYVE28*, *ADAMTS18* and *CERS4*.

Functional characterization provided new evidence for the regulatory effects of the representative non-coding variants, particularly rs4977199 (*BOP1*), rs2303115 (*STXBP2*) and rs2927712 (*CERS4*), on modulating the activities of enhancers, transcription, and transcription factors, respectively (Table 4). The functional predictions also revealed thirty-six previously uncharacterized genes functioning in host defence, leukocyte migration, and the cellular responses to interferon-gamma (Figure 2), and highlighted the four genes— *UQCRFS1*, *PTPN6*, *RALY* and *ZMYM4*—associated with aging, forebrain and nervous system morphology, lung and bone morphology, and neurodegenerative diseases and blood disorders, respectively, among other traits.

Across ancestries, the most striking result is represented by the association of rs10196189 (*GALNT13*) with sprinting ability in populations of West African, East Asian and European descent, crossing the GWAS significance threshold of *P* = 5E-08 (Table 3). Gene-based analysis unveiled a statistically significant association for *ZFYVE28*, followed by *ADAMTS18* at the borderline significance, across West Africans and East Asians. All three genes are universally expressed in the nervous tissue system (Figure 2 and Supplementary Fig. S4). For cohort-specific findings, multiple lines of evidence are converged on the findings: *MPRIP* (T-allele of rs117143557; Jpn), *BOP1* (G-allele of rs4977199; Jam), *GRM7* (A-allele of rs2875287; A-A), *CERS4* (C-allele of rs2927712; Jam and A-A), and *STXBP2* (A-allele of rs2303115; Jam) (Table 4); according to the strength of SNP- and gene-associations and/or their regulatory effect scores in Sei. Intriguingly, rs4977199, rs2927712 and rs2303115 are statistically significant *cis*-eQTLs for several other genes in blood-derived expression^36^, such as rs4977199 for *DGAT1* (encoding a key metabolic enzyme), rs2927712 for *CD320* (involved in B-cell proliferation) and rs2303115 for *PCP2* (predicted to locate in neuronal cell body). It is worth noting the large effect of the T-allele of rs117143557 (MAF > 5%) nearest *MPRIP* on elite sprinting ability in Japanese athletes (Table 4) while being present in a much lesser content in European and African populations (MAF < 1%); indicative of a potential population-specific function of the variant. These genes are also specifically expressed in the nervous tissues system, except for *MPRIP* (in multiple tissue systems, including the nervous tissues) and *STXBP2* (in the hematopoietic system) (Figure 2 and Supplementary Fig. S4). In contrast to the SNP- and gene-based analyses supporting the role of *GALNT13*, *MPRIP* and *GRM7* in this study, the lack of functional evidence available for them in Sei underscores the novelty of these associations and highlights an opportunity to characterize their functions with reference to human performance, fitness, and health.

Tissue-specific network analysis for the putative genes above provides further exploratory links focusing on their most relevant interacting genes in specific tissues and biological processes, detailed in the Functional networks (GIANT) section. These predicated and functionally similar genes can be populated into the wider biological contexts for reforming and testing the hypotheses for their roles in the nervous system, forebrain development, glutamate signalling pathway, higher neurological functions in caudate nucleus and leukocyte migration (Supplementary Fig. S4).

Other notable findings from this study include the identification of 36 novel genes in populations of West Africans (Jam and A-A) and East Asians (Jpn) associated with the specific functional modules indicative of response to virus (M1), viral life cycle (M2), cell apoptosis (M3), response to interferon-gamma (M4) and leukocyte migration (M5) (Figure 3). These results coincide with the recent discoveries of *IFI44*, *OAS2*, and *HERC6* (present in M1) being present in IFN responsive T cells in SARS-CoV-2 infected macaques in suppressing viral transcription and aiding clearance of the virus^37^. *IFI44* has also previously been reported to repress HIV-1 replication *in vitro*^38^. Evidence for *NMI* and *IFI35* (present in M2) showed that they act as damage-associated molecular patterns in the extracelluar space for their proinflammatory functions in cellular infection and damage through activating the nuclear factor-κB in the Toll-like receptor 4 signalling pathway^39^; more recently, *IFI35* has been proposed as a biomarker of neuroinflammation and for predicting the long-term treatment response in multiple sclerosis patients^40^. Autosomal recessive mutations in *IL2RB* (present in M5) have only been recently characterized in severe autoimmunity and viral susceptibility, reflecting the important functions of *IL2RB* in T and NK cells signalling and in maintaining immune tolerance^41^. Conceivably, the functional module-specific genes uncovered here provide new insights for future studies aiming to understand how these molecular factors within and between the modules could shape immunity, thereby informing the development of therapeutics for multisystem autoimmune diseases and cancer particularly in understudied populations. Intriguingly, we have also been able to reprioritize four genes with respect to their functional significance derived from common genetic variations in Jam (*UQCRFS1*), A-A (*PTPN6*), Jpn (*RALY*), and across Jam, A-A and Jpn (*ZMYM4*), in addition to mutations in these genes causing mitochondrial disorders (manifesting in severe multi-systemic disorders^42^), inflammatory diseases^43^, metabolic disease states^44^, and energy dyshomeostasis^45^, respectively. These results provide an interface for closely studying any connections between common regulatory variants and rarer protein-coding mutations in these genes in human health and disease.

Taken together, this multi-ethnic, multi-phase GWAS has allowed us to unequivocally identify nine genes and their representative SNPs (where applicable) in association with world-class sprinting ability in an ancestry-consistent and an ancestry-specific manner. We have also been able to elaborate on their functional roles and interacting genes in a tissue-/cell-type-specific context, illustrate thirty-six novel genes functioning in cellular immune responses, and highlight four genes in connection with aging as well as neurological, blood and bone disorders from the elite athletic performance cohorts. It is noteworthy that this work focused on understudied ancestral groups from West Africa and East Asia comprising athletes of the highest performance calibre, thereby reflecting the highest quality of extreme phenotypes studied here and adding further values to the current genomic landscape lacking genetic diversity. Overall, our results provide new biological insights into elite sprint performance and open new avenues for future research of personalized exercise prescription and precision medicine.

## Methods

### Sample collection

#### Elite Jamaican and African-American sprint cohorts (of West African ancestry)

These cohorts comprise Jamaican and African-American sprint athletes of the highest caliber and geographically matched controls. One hundred and sixteen Jamaican athletes (male = 60, female = 56) and 311 matched controls (male = 156, female = 155) were recruited^46^. Among them, 71 and 35 participated in the 100 – 200 m and 400 m sprint events, respectively, and 10 were involved in the jump and throw events. They can be further classified into national-(n = 28) and international-level (n = 88) athletes, representing Jamaica and the Caribbean at major international competitions. Among the 88 international-level athletes, 46 won medals at the international events or held world records in sprinting. Subject to DNA availability and quality, 95 athletes and 102 controls were included in the current study.

One hundred and fourteen African-American sprint athletes (male = 62, female = 52) and 191 matched controls (male = 72, female = 119) were recruited^46^. Among the athletes, 48, 42 and 24 participated in the 100-200 m, 400 m, and jump and throw events, respectively. Athletes can be sub-divided into 28 national- and 86 international-level athletes; 35 of these athletes won medals at international games or broke sprint world records. Subject to DNA availability and quality, 108 athletes and 47 controls were included in the current study. An additional 350 African-American controls were also included, outsourced from published data^47^; to boost the number of African-American controls for this analysis.

#### Elite Japanese sprint athlete cohort (of East Asian ancestry)

This cohort involved 54 (male = 48, female = 6) international-level sprint athletes in Japan, but they were not necessarily medallists. One hundred and eighteen geographically matched controls (male = 38, female = 80) were recruited from the general population.

#### Elite European athlete cohorts

Cohort 1 comprises 133 sprinters, 234 power-oriented athletes, 451 endurance athletes and 1,525 controls from Belarus, Lithuania, and Russia. Cohort 2 consists of 171 sprinters, 168 power-oriented athletes, 252 endurance athletes and 595 controls from Australia, Belgium, Greece, and Poland. All athletes are national-, and international-level, or are prize winners of these events.

Ethics Committee of the University of West Indies, Jamaica (protocol number: ECP 121, 2006/2007), the Institutional Review Board of Florida State University, USA (D5.158), the Institutional Review Board of Juntendo University, Japan (SHSS 2022-137), the Institutional Review Board of Tokyo Metropolitan Institute of Gerontology, Japan (TMIG 19-3784), the Institutional Review Board of the National Institute of Health and Nutrition, Japan (KENEI 2-09) Japan, the Bioethics Commission of Institute of Bioorganic Chemistry of the National Academy of Sciences of the Republic of Belarus, Belarus (2017/03), the Lithuanian Bioethics Committee, Lithuania (69-99-111), the Ethics Committee of the Federal Research and Clinical Center of Physical-Chemical Medicine of the Federal Medical and Biological Agency of Russia, Russia (2017/04), the Institutional Review Board of the Children’s Hospital at Westmead, Australia (2003/086), the Royal Children’s Hospital Human Research Ethics Committee, Australia (35172), the Ethical Committee of Ghent University Hospital, Belgium (B67020097348), Aristotle University of Thessaloniki Research Committee, Greece (1895), and the Pomeranian Medical University Ethics Committee, Poland (BN-001/45/08) approved for this work. Written informed consent was obtained from all subjects. All research was performed in accordance with the relevant guidelines and regulations, and in accordance with the “Declaration of Helsinki”.

### DNA processing for GWAS

DNA was isolated from buccal cells or whole saliva. Participants were asked not to consume food or drink for at least 30 minutes before providing a sample. Buccal cells were collected by a trained individual by firmly rubbing a brush (Medical Packaging Corporation, Camarillo, CA, USA) against the inside of a participant’s cheek for at least 15 seconds. The head of the brush was then cut into a screw cap tube containing cell lysis solution (0.1 M Tris-HCl pH 8.0, 0.1 M EDTA; 1 % SDS). DNA was extracted using the QIAamp DNA Mini kit (QIAgen, Hilden, Germany) according to the manufacturer’s instructions with minor adjustments.

Briefly, 500 mL of the sample was transferred to a clean 1.5 mL microcentrifuge tube, followed by adding in 15 µL proteinase K and incubated at 55°C for 30 minutes to 1 hour in an air incubator (Binder B28, BINDER GmbH, Tuttlingen, Germany). Absolute ethanol (500 mL) was then added to the sample, vortexed thoroughly and the mixture was transferred to a spin column for centrifugation. DNA was washed twice using 500 mL of AW1 and AW2 buffers and was dissolved in 200 mL of AE buffer (10 mM Tris-Cl; 0.5mM EDTA; pH 9.0). Whole saliva was collected using the Oragene DNA Self Collection Kit (OG-250, DNA Genotek Inc., Canada). About 2 mL of saliva was collected and sufficiently mixed with the Oragene chemistry (DNA Genotek Inc., Canada) by repeated inversion for 10 seconds. DNA was extracted following the manual purification of DNA from 0.5 mL of sample using DNA Genotek’s prepIT·L2P DNA extraction kit with minor adjustments. In brief, 500 mL of the saliva sample was transferred into a 1.5 mL microcentrifuge tube. Twenty microliter of the Oragene DNA purifier solution was then added and mixed by vortexing for 3 seconds. The mixture was placed on ice for 10 minutes before centrifugation at room temperature for 10 minutes at 13,000 rpm (15,000 × *g*). The supernatant was then transferred into a fresh tube and an equal volume of absolute ethanol (500 mL) was added, mixed by inverting gently 10 times. The mix was incubated at room temperature for 10 minutes to allow full precipitation of DNA, followed by centrifugation at room temperature for 2 minutes at 13,000 rpm. The supernatant was then removed and discarded, and the pellet was dried in an air incubator at 50°C for about 20 minutes and was dissolved in 300 mL of TE buffer (100 mM Tris, 10 mM EDTA, pH 8.0), allowing complete rehydration of DNA overnight at room temperature. DNA was quantified using the Nanodrop Technologies Nanodrop ND-8000 Spectrophotometer (Wilmington, DE, USA).

Purified DNA samples from the Jamaican and African-American cohorts were shipped to Tokyo Metropolitan Institute of Gerontology, Japan, for genotyping on HumanOmniExpress and HumanOmni1-Quad Beadchips (Illumina, San Diego, California, USA). During transportation, DNA was stored in the sterile Thermo Scientific Matrix Storage Tubes (0.75 mL, 8 x 12 format; Thermo Fisher Scientific, Hudson, New Hampshire, USA) and was shipped with dry ice. DNA quality was reevaluated using the PicoGreen Assay prior to the whole-genome genotyping; and only samples of at least 50 ng of DNA were taken forward.

### Discovery phase – part one: GWAS data analysis workflow

Illumina GenomeStudio Software (v2010.3), PLINK^48^, EIGENSTRAT^49^, were used for converting array outputs to PLINK formats, genotype QCs and association testing, and population stratification analyses, respectively.

Per-individual and per-marker QCs were initially performed in a cohort-specific manner. SNPs met the following inclusion criteria for inclusion: maximal proportion of missing SNPs per sample of 5% (267,969 Jam; 297,404 A-A; 48,490 Jpn; SNPs excluded); MAF of 1% (9 A-A; SNPs excluded); minimum Hardy-Weinberg disequilibrium frequency *P*-value 1 x 10^-7^ (31 Jpn; SNPs excluded). Any samples failing per-sample QCs were removed, including discordant sex, low call rate below 95%, heterozygosity rate exceeding three standard deviations from mean heterozygosity, cryptic relatedness subject to proportion IBD (identity-by-descent) of 0.05 and visual inspection of the relationship between Z0 and Z1 values, and outliers of principal component analysis (Supplementary Table S3, and Supplementary Fig. S10 and S11). Genetic associations were evaluated using logistic regression for 22 autosomes in PLINK, assuming an additive effect in Jam and A-A and taking into account the top 10 principle components and genotyping centre effect in the model, where appropriate. Standard allelic association analysis was performed in PLINK, comparing the allele-frequency differences in Jpn. Visualisation of the GWAS results and regional associations was carried out in the Bioconductor package “karyoloteR”^50^ and the LocusZoom^51^, respectively; along with other packages, “Cairo”^52^, “tidyr”^53^, and “cpvSNP”^54^, required for plotting.

### Discovery phase – part two: imputation and meta-analyses

Two imputation workflows were adopted: IMPUTE2 (phasing with SHAPEIT2^55^) on the 1000G phase 3 reference panel^25,56^ and the Sanger Imputation Server on the African Genome Resources^26^ (phasing with EAGLE2^57^, imputing with PBWT^58^). Post-imputation QCs included imputation quality measure > 0.3 and MAF ≥ 5%. Cohort-specific genetic associations were then tested on the SNPTEST^59^ using a frequentist additive model, conditioning on the top 10 principle components (accounting for population structure) and genotyping centres, where appropriate. Meta-analyses using the inverse variance-weighted fixed-effect model of the three imputed cohorts were performed in METAL^60^ subjected to genomic control (GC) correction.

### Replication phase: independent replications of rs10196189 in athletes of European descent

Genotyping was conducted for rs10196189 in the two independent European cohorts (Cohort 1 and Cohort 2) of sprint and power-oriented athletes, endurance-oriented athletes, and matched controls. Genetic associations were tested on both additive and dominant genetic models for each replication cohort. METAL^60^ was then used to perform a meta-analysis of the discovery and replication findings for rs10196189 across the three ancestral populations (West-Africans, East-Asians and Europeans).

### Gene analysis phase – part one: gene- and gene-set enrichment analyses

Gene analysis of the Sanger imputation/SNPTEST summary statistics was performed using MAGMA v1.09b^61^. SNPs were annotated to NCBI 37.3 and assigned to a gene if located within 5kb up-or downstream of the gene region. The “multi” gene analysis model was applied, testing both “mean” and “top” SNP associations, which are sensitive to detect associations in high LD regions of a gene using the sum of squared SNP *Z*-statistics as the test statistic or in the top proportion of SNPs using the lowest SNP *P*-value as the test statistic. Gene location files derived from NCBI 37.3 and LD estimations between SNPs using the African and East Asian reference populations of the 1000 Genomes Project were downloaded from the auxiliary files available at https://ctg.cncr.nl/software/magma. Fixed effects meta-analysis of the gene analysis results in METAL was conducted across Jam, A-A and Jpn, followed by competitive gene-set analysis using the MSigDB (v7.4) hallmark^28^ (N=50) and canonical pathways (C2:CP; N=2,922) collections of functional gene sets. Multiple testing correction was applied according to the number of genes available for each analysis.

### Gene analysis phase – part two: gene-based heritability estimation

Gene-based heritability was estimated using individual-level data following the Sanger imputation/SNPTEST analyses by restricted maximum likelihood (REML^62^) test implemented in LDAK5.2^63^. Again, SNPs were annotated to NCBI 37.3 and assigned to a gene if located within 5kb up-or downstream of the gene region. SNPs were scaled to the recommended power of −0.25, implying weak negative selection (See: dougspeed.com/gene-based-analysis/). Covariates were included to correct for population structure and genotyping centre effect, where appropriate. Heritability estimates were reported on both observed and liability scales by setting the population prevalence to 1% in the latter. This was based off a conservative estimate from estimated probabilities of competing in College Athletics from the US Federal associations, the National Federation of State High School Associations and the National Collegiate Athletic Association (NCAA)^64^ (i.e., 2.8% and 1.9% in women and men respectively for Track and Field in NCAA Division I in 2018-19). Estimates for competing in professional athletics or at the Olympic level for Track and Field were not available, but this is believed to be only a select few (well under 1%). Here we set an arbitrary liability scale to 1%, only as an indication of low prevalence.

### Final phase of data-driven prediction for tissue-specific gene functions, regulation and GWAS re-prioritization in humanbase

To functionally annotate findings from both SNP- and gene-based analyses, we utilised humanbase (https://humanbase.io/). Humanbase hosts a suite of machine-learning algorithms (Naïve Bayes, deep learning, and Supporter Vector Machine) to enable functional predictions of genomic variants and genes. By integrating 987 genomic datasets, consisting of ∼38,000 conditions from ∼14,000 publications, and by accessing their relevance in 144 tissues and cell lineages, it provides a high throughput and unbiased method to prioritize genes and genetic variations for follow-up. Here, we applied tissue-specific functional network (GIANT^29^) for the top genes identified from the gene-based analyses including *GALNT13*, *STXBP2*, *BOP1*, *HSF1*, *GRM7*, *MPRIP*, *ZFYVE28* and *ADAMTS18* and the Sei analysis^31^ for six sentinel SNPs representing independent regional signals resided near or in *GALNT13* (rs113303758, T/C), *STXBP2* (rs2303115, G/A), *BOP1* (rs4977199, T/G), *GRM7* (rs3864067, A/G), *MPRIP* (rs117143557, C/T), and *CERS4* (rs2927712, A/C) for tissue-/cell-type-specific regulatory activities. We have also performed functional module^30^ detection in blood for the gene-set of interferon-gamma response encompassing 199 genes resulted from the gene-set enrichment analysis from Magma; to investigate genetic associations with specific biological processes represented in cohesive gene clusters or functional modules, including potential genetic associations depicted by functionally uncharacterized genes in the detected modules.

Lastly, we conducted NetWAS^29^ analysis for nominally significant genes (*P* < 0.01) derived from the gene-based analysis in a relevant tissue network representative of the top gene finding in each cohort. Specifically, 18,416 (Jam), 18,414 (A-A), and 18,317 (Jpn) and 13,301 (meta-analysis) genes were uploaded for gene reprioritization in the central nervous system, forebrain, brain, and caudate nucleus, respectively. The resulting positive NetWAS scores indicate genes that are more likely to be nominally significant. To evaluate the reprioritized gene lists further, we submitted them to Enrichr^33–35^ for functional annotations focusing on gene enrichment in “Diseases/Drugs” and “Cell Types” gene-set libraries.

### Statistical analysis

Genomic data were analyzed at both SNP- and gene-level across the genome, using the specific software and packages detailed in the sections above. For the GWAS association results, a *P* value of 5E-05 served as a suggestive cut-off. For the imputed and meta-analyzed data datasets, *P* < 1E-05 and *P* < 5E-08 were used to filter the results. For the gene- and gene-set enrichment analysis, Bonferroni or Benjamini–Hochberg (BH) corrections were applied to correct for multiple tests; only statistically significant associations or borderline significance were reported. For gene-based heritability estimation, the *P* value is calibrated using 10 permutations (default setting in LDAK5.2). For the GIANT analysis, enriched biological processes and pathways were declared significant at *P* < 0.05. For the functional module analysis, the Q value of each term is calculated using the one-sided Fisher’s exact test and BH corrections for multiple tests (implemented in humanbase). The Enrichr Q value is an adjusted *P* value using the BH method.

## Consortium

### Sportgene Research Group

Emiliya S. Egorova^9^, Leysan J. Gabdrakhmanova^9^, Ekaterina A. Semenova^11,26^, Nikolay A. Kulemin^11^, Andrey K. Larin^11^, Irina V. Haidukevich^14^, Irina L. Gilep^27^, Egor B. Akimov^28^

^26^Volga Region State University of Physical Culture, Sport and Tourism, Kazan, Russian Federation.

^27^Republican Scientific and Practical Center of Sports, Minsk, Belarus.

^28^Meret Solutions Mental Health Association, Madison, United States.

## Data Availability

Access to all the elite athletic performance cohorts may be limited by participant consent and data sharing agreements; requests should be directed in the first instance via the corresponding authors. All other data are available in the main text or the supplementary materials.

## Acknowledgements

We would like to thank all participants of this project. We would also like to thank Dr. Olga N. Pyankova for her assistance in performing the genotyping replication of the European Cohort 1. The GWAS genotyping work was funded by JSPS KAKENHI grant no. 15H03081 and 22H03486 to N.F. Genotyping replication in the Belorussian samples was funded by the framework of the Program for Basic Research in the Russian Federation for a long-term period (2021–2030) grant no. 122030100168-2 to A.A.G. The additional 350 African American controls were drawn from the Genetics of Early Onset Stroke (GEOS) Study, with funding support from NIH U01 HG-004436 to B.D.M.

## Author contributions

Conceptualization: G.W., N.F., Y.P.P.; formal analysis: G.W.; funding acquisition: N.F., A.A.G., B.D.M.; methodology: G.W., N.F., E.M.M., H.M., F.C.G., S.P., Y.P.P.; resources: G.W., Y.C.C., B.D.M., E.M., K.G.A., S.P., Y.P.P.; supervision: S.P., Y.P.P.; replication: I.I.A., Sportgene Research Group, E.V.G., M.L.F., A.A.G., V.G., C.N.M., T.V., P.C., W.D., I.P., F.C.G., K.N.; visualization: G.W.; writing-original draft: G.W.; writing-review and editing: G.W., N.F., E.M.M., M.T., M.M., H.M., Y.C.C., B.D.M., E.M., K.G.A., I.I.A., Sportgene Research Group, E.V.G., M.L.F., A.A.G.,V.G., C.N.M., T.V., P.C., W.D., I.P., F.C.G., K.N., S.P., Y.P.P.

## References

1 Rankinen, T. et al. The human gene map for performance and health-related fitness phenotypes. Med Sci Sports Exerc 33, 855–867 (2001).

2 Rankinen, T. et al. The human gene map for performance and health-related fitness phenotypes: the 2001 update. Med Sci Sports Exerc 34, 1219–1233 (2002).

3 Pérusse, L. et al. The human gene map for performance and health-related fitness phenotypes: the 2002 update. Med Sci Sports Exerc 35, 1248–1264 (2003).

4 Rankinen, T. et al. The human gene map for performance and health-related fitness phenotypes: the 2003 update. Med Sci Sports Exerc 36, 1451–1469 (2004).

5 Wolfarth, B. et al. The human gene map for performance and health-related fitness phenotypes: the 2004 update. Med Sci Sports Exerc 37, 881–903 (2005).

6 Rankinen, T. et al. The human gene map for performance and health-related fitness phenotypes: the 2005 update. Med Sci Sports Exerc 38, 1863–1888 (2006).

7 Bray, M.S. et al. The human gene map for performance and health-related fitness phenotypes: the 2006-2007 update. Med Sci Sports Exerc 41, 35–73 (2009).

8 Rankinen, T. et al. Advances in exercise, fitness, and performance genomics. Med Sci Sports Exerc 42, 835–846 (2010).

9 Hagberg, J.M. et al. Advances in exercise, fitness, and performance genomics in 2010. Med Sci Sports Exerc 43, 743–752 (2011).

10 Roth, S.M. et al. Advances in exercise, fitness, and performance genomics in 2011. Med Sci Sports Exerc 44, 809–817 (2012).

11 Pérusse, L. et al. Advances in exercise, fitness, and performance genomics in 2012. Med Sci Sports Exerc 45, 824–831 (2013).

12 Wolfarth, B. et al. Advances in exercise, fitness, and performance genomics in 2013. Med Sci Sports Exerc 46, 851–859 (2014).

13 Loos, R.J. et al. Advances in exercise, fitness, and performance genomics in 2014. Med Sci Sports Exerc 47, 1105–1112 (2015).

14 Sarzynski, M.A., et al. Advances in Exercise, Fitness, and Performance Genomics in 2015. Med Sci Sports Exerc 48, 1906–1916 (2016).

15 Willems, S.M. et al. Large-scale GWAS identifies multiple loci for hand grip strength providing biological insights into muscular fitness. Nat Commun 8, 16015 (2017).

16 Nakamichi, R. et al. The mechanosensitive ion channel PIEZO1 is expressed in tendons and regulates physical performance. Sci Transl Med 14, eabj5557 (2022).

17 Tabor, H.K., Risch, N.J., Myers, R.M. Candidate-gene approaches for studying complex genetic traits: practical considerations. Nat Rev Genet 3, 391–397 (2002).

18 Shendure, J. et al. DNA sequencing at 40: past, present and future. Nature 550, 345–353 (2017).

19 Tishkoff, S.A. et al. The genetic structure and history of Africans and African Americans. Science 324, 1035–1044 (2009).

20 Sirugo, G., Williams, S.M., Tishkoff, S.A. The Missing Diversity in Human Genetic Studies. Cell 177, 26–31 (2019).

21 Peterson, R.E. et al. Genome-wide Association Studies in Ancestrally Diverse Populations: Opportunities, Methods, Pitfalls, and Recommendations. Cell 179, 589–603 (2019).

22 Hong, E.P., Park, J.W. Sample size and statistical power calculation in genetic association studies. Genomics Inform 10, 117–122 (2012).

23 Uffelmann, E., et al. Genome-wide association studies. Nature Reviews Methods Primers 1, 59 (2021).

24 Klein, R.J. et al. Complement factor H polymorphism in age-related macular degeneration. Science 308, 385–389 (2005).

25 Howie, B.N., Donnelly, P., Marchini, J. A flexible and accurate genotype imputation method for the next generation of genome-wide association studies. PLoS Genet 5, e1000529 (2009).

26 McCarthy, S. et al. A reference panel of 64,976 haplotypes for genotype imputation. Nat Genet 48, 1279–1283 (2016).

27 ENCODE Project Consortium. An integrated encyclopedia of DNA elements in the human genome. Nature 489, 57–74 (2012).

28 Liberzon, A. et al. The Molecular Signatures Database (MSigDB) hallmark gene set collection. Cell Syst 1, 417–425 (2015).

29 Greene, C.S. et al. Understanding multicellular function and disease with human tissue-specific networks. Nat Genet 47, 569–576 (2015).

30 Krishnan, A. et al. Genome-wide prediction and functional characterization of the genetic basis of autism spectrum disorder. Nat Neurosci 19, 1454–1462 (2016).

31 Chen, K.M., Wong, A.K., Troyanskaya, O.G., Zhou, J. A sequence-based global map of regulatory activity for deciphering human genetics. Nat Genet 54, 940–949 (2022).

32 Wang, Q. et al. AtHDA6 functions as an H3K18ac eraser to maintain pericentromeric CHG methylation in Arabidopsis thaliana. Nucleic Acids Res 49, 9755–9767 (2021).

33 Chen, E.Y. et al. Enrichr: interactive and collaborative HTML5 gene list enrichment analysis tool. BMC Bioinformatics 14, 128 (2013).

34 Kuleshov, M.V. et al. Enrichr: a comprehensive gene set enrichment analysis web server 2016 update. Nucleic Acids Res 44, W90–97 (2016).

35 Xie, Z. et al. Gene Set Knowledge Discovery with Enrichr. Curr Protoc 1, e90 (2021).

36 Võsa, U. et al. Large-scale cis-and trans-eQTL analyses identify thousands of genetic loci and polygenic scores that regulate blood gene expression. Nat Genet 53, 1300–1310 (2021).

37 Singh, D.K. et al. Myeloid cell interferon responses correlate with clearance of SARS-CoV-2. Nat Commun 13, 679 (2022).

38 Power, D. et al. IFI44 suppresses HIV-1 LTR promoter activity and facilitates its latency. Virology 481, 142–150 (2015).

39 Xiahou, Z. et al. NMI and IFP35 serve as proinflammatory DAMPs during cellular infection and injury. Nat Commun 8, 950 (2017).

40 De Masi, R., Orlando, S. IFI35 as a biomolecular marker of neuroinflammation and treatment response in multiple sclerosis. Life Sci 259, 118233 (2020).

41 Campbell, T.M., Bryceson, Y.T. IL2RB maintains immune harmony. J Exp Med 216, 1231–1233 (2019).

42 Gusic, M. et al. Bi-Allelic UQCRFS1 Variants Are Associated with Mitochondrial Complex III Deficiency, Cardiomyopathy, and Alopecia Totalis. Am J Hum Genet 106, 102–111 (2020).

43 Speir, M. et al. Ptpn6 inhibits caspase-8-and Ripk3/Mlkl-dependent inflammation. Nat Immunol 21, 54–64 (2020).

44 Zhang, Z. et al. Collaborative interactions of heterogenous ribonucleoproteins contribute to transcriptional regulation of sterol metabolism in mice. Nat Commun 11, 984 (2020).

45 Marenne, G. et al. Exome Sequencing Identifies Genes and Gene Sets Contributing to Severe Childhood Obesity, Linking PHIP Variants to Repressed POMC Transcription. Cell Metab 31, 1107–1119.e1112 (2020).

46 Scott, R.A. et al. ACTN3 and ACE genotypes in elite Jamaican and US sprinters. Med Sci Sports Exerc 42, 107–112 (2010).

47 Cheng, Y.C. et al. Genome-wide association analysis of ischemic stroke in young adults. G3 (Bethesda) 1, 505-514 (2011).

48 Purcell, S. et al. PLINK: a tool set for whole-genome association and population-based linkage analyses. Am J Hum Genet 81, 559–575 (2007).

49 Price, A.L. et al. Principal components analysis corrects for stratification in genome-wide association studies. Nat Genet 38, 904–909 (2006).

50 Gel, B., Serra, E. karyoploteR: an R/Bioconductor package to plot customizable genomes displaying arbitrary data. Bioinformatics 33, 3088–3090 (2017).

51 Pruim, R.J. et al. LocusZoom: regional visualization of genome-wide association scan results. Bioinformatics 26, 2336–2337 (2010).

52 Urbanek, S., Horner, J. R Graphics Device using Cairo Graphics Library for Creating High-Quality Bitmap (PNG, JPEG, TIFF), Vector (PDF, SVG, PostScript) and Display (X11 and Win32) Output, <https://cran.r-project.org/web/packages/Cairo/Cairo.pdf> (2022).

53. Wickham, H., Girlich, M. tidyr: Tidy Messy Data <https://tidyr.tidyverse.org, https://github.com/tidyverse/tidyr.> (2022).

54 McHugh, C., Larson, J., Hackney, J. cpvSNP: Gene set analysis methods for SNP association p-values that lie in genes in given gene sets. R package version 1.30.0., 2022).

55 Delaneau, O., Marchini, J. Integrating sequence and array data to create an improved 1000 Genomes Project haplotype reference panel. Nat Commun 5, 3934 (2014).

56 Howie, B., Marchini, J., Stephens, M. Genotype imputation with thousands of genomes. G3(Bethesda) 1, 457-470 (2011).

57 Loh, P.R. et al. Reference-based phasing using the Haplotype Reference Consortium panel. Nat Genet 48, 1443–1448 (2016).

58 Durbin, R. Efficient haplotype matching and storage using the positional Burrows-Wheeler transform (PBWT). Bioinformatics 30, 1266–1272 (2014).

59 Marchini, J., Howie, B. Genotype imputation for genome-wide association studies. Nat Rev Genet 11, 499–511 (2010).

60 Willer, C.J., Li, Y., Abecasis, G.R. METAL: fast and efficient meta-analysis of genomewide association scans. Bioinformatics 26, 2190–2191 (2010).

61. de Leeuw, C.A., Mooij, J.M., Heskes, T., Posthuma, D. MAGMA: generalized gene-set analysis of GWAS data. PLoS Comput Biol 11, e1004219 (2015).

62 Speed, D., Cai, N., Johnson, M.R., Nejentsev, S., Balding, D.J. Reevaluation of SNP heritability in complex human traits. Nat Genet 49, 986–992 (2017).

63 Zhang, Q., Privé, F., Vilhjálmsson, B., Speed, D. Improved genetic prediction of complex traits from individual-level data or summary statistics. Nat Commun 12, 4192 (2021).

64 National Collegiate Athletic Association. Estimated Probability of Competing in College Athletics, <https://ncaaorg.s3.amazonaws.com/research/pro_beyond/2020RES_ProbabilityBeyondHSFiguresMethod.pdf> (2020).

