## Supplementary Figures and Tables for "Multi-phase, multi-ethnic GWAS uncovers putative loci in predisposition to human sprint performance, health and disease"

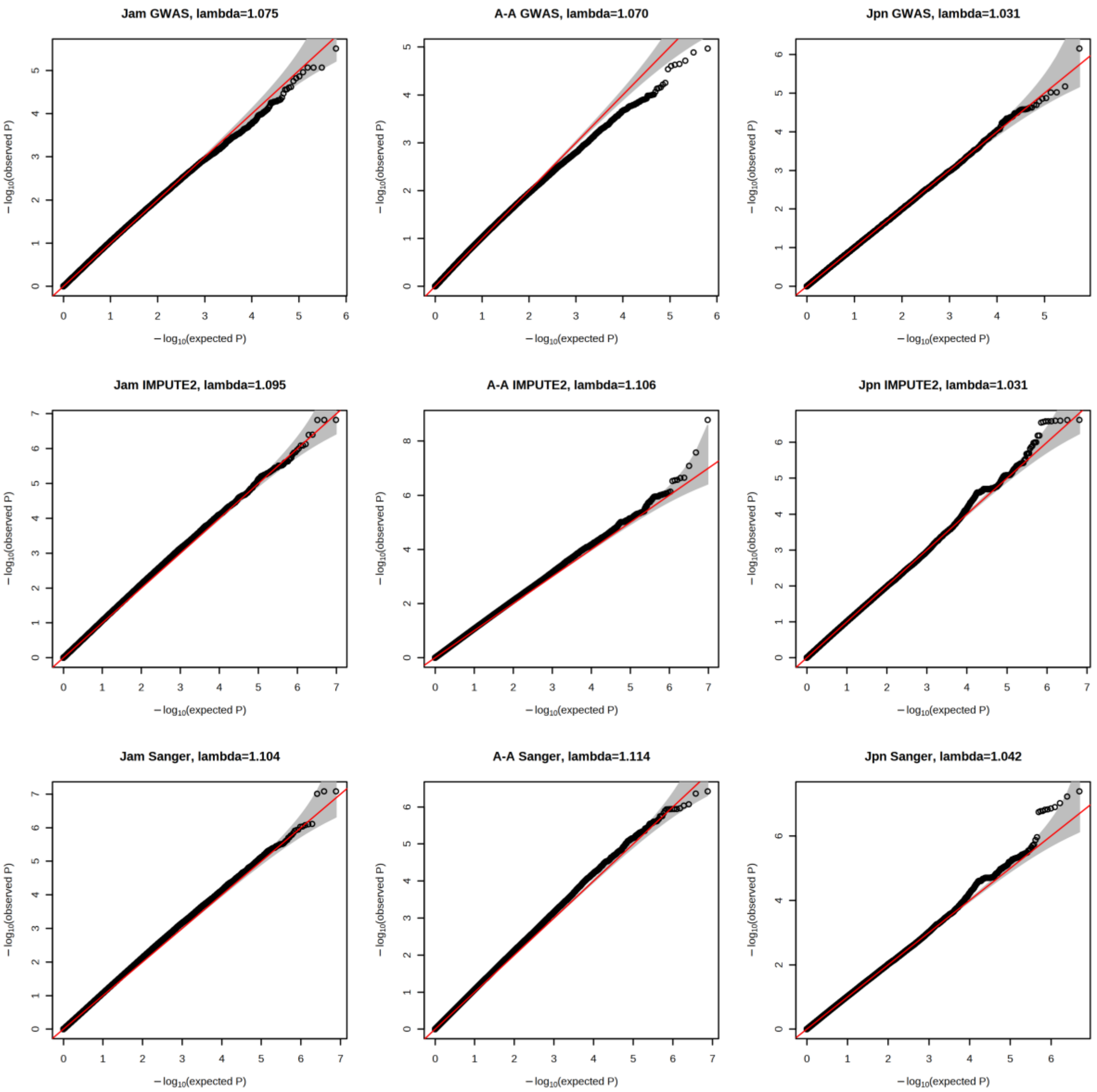


Supplementary Fig. S1. Q-Q plots of the GWAS, IMPUTE2 and Sanger Imputation Server results in Jam, A-A, and Jpn cohorts.

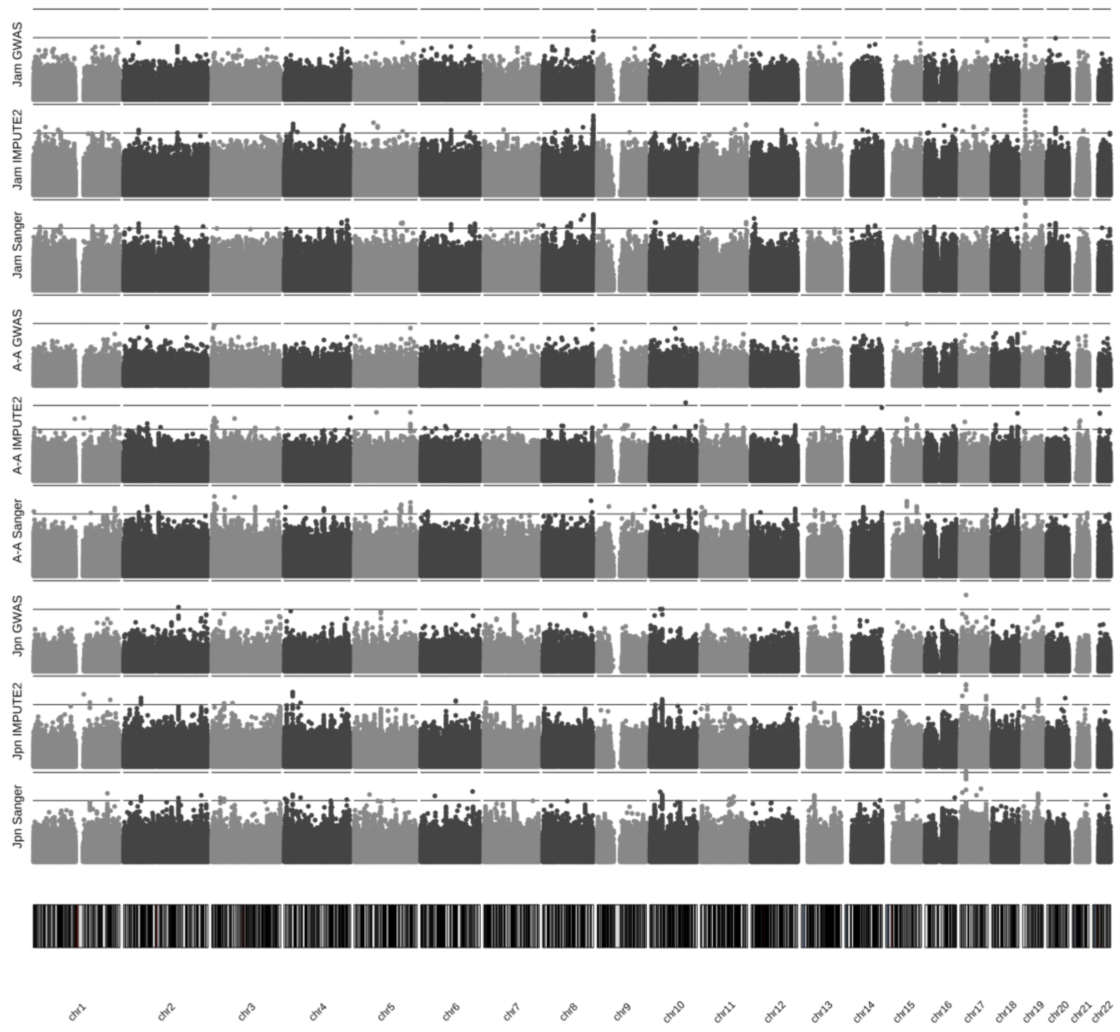


Supplementary Fig. S2. Manhattan plots of the GWAS, IMPUTE2 and Sanger Imputation Server results in Jam, A-A, and Jpn cohorts across 22 autosomes. Dotted line: suggestive significance cut-off of 1E-05; dashed line: genome-wide significance cut-off of 5E-08.

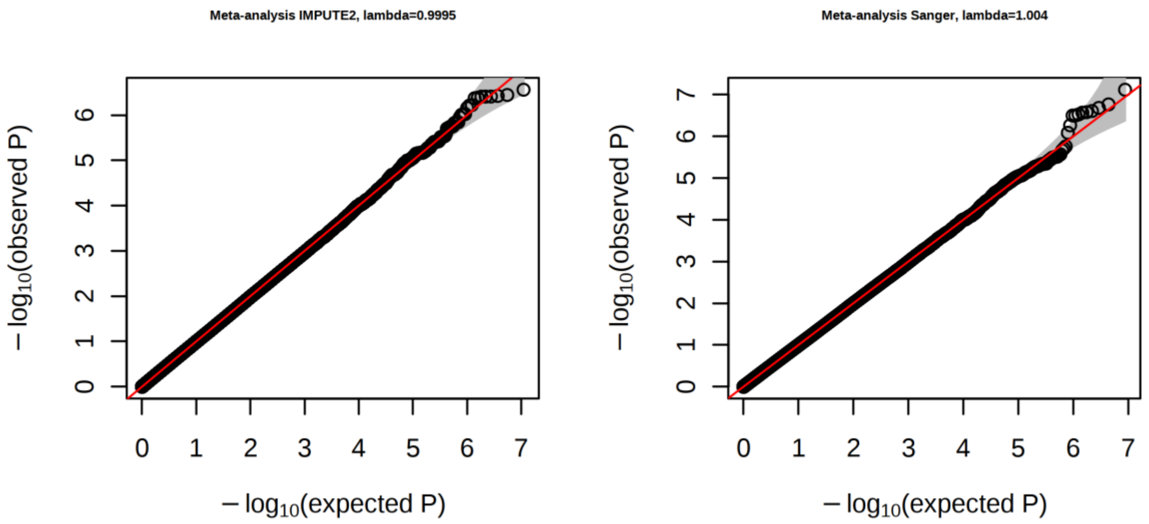


Supplementary Fig. S3. QQ-plots of the meta-analysis of the imputed results across Jam, A-A and Jpn, following imputation using IMPUTE2 and Sanger Imputation server.

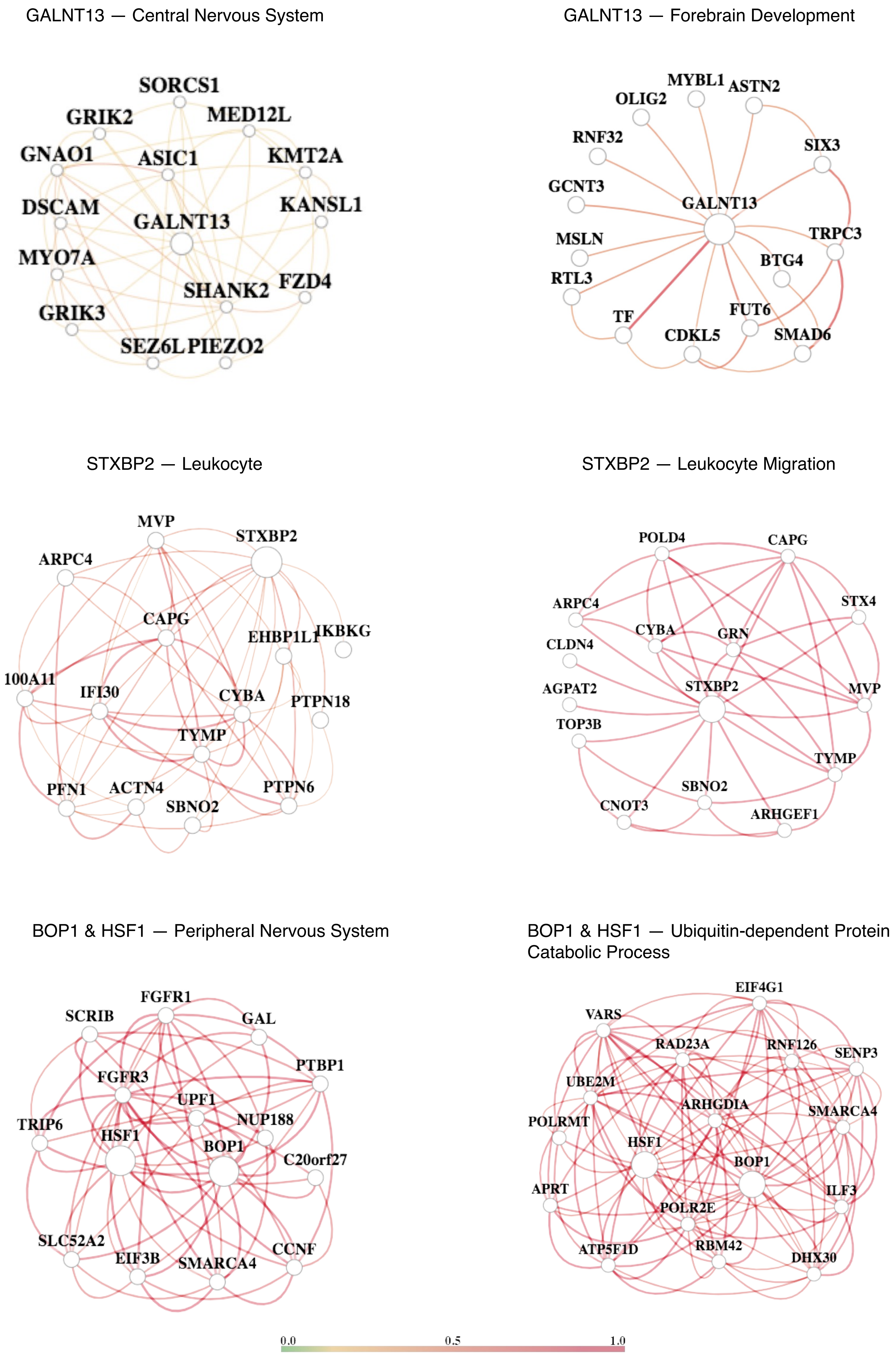
GALNT13 — Central Nervous System
SORCS1
GRIK2
MED12L
GNAO1
ASIC1
KMT2A
DSCAM
KANSL1
GALNT13
MYO7A
SHANK2
FZD4
GRIK3
SEZ6L
PIEZO2
GALNT13 — Forebrain Development
MYBL1
OLIG2
ASTN2
RNF32
SIX3
GCNT3
GALNT13
MSLN
TRPC3
BTG4
RTL3
FUT6
TF
CDKL5
SMAD6
STXBP2 — Leukocyte
MVP
STXBP2
ARPC4
CAPG
EHBP1L1
IKBKG
100A11
IFI30
CYBA
PTPN18
TYMP
PFN1
ACTN4
PTPN6
SBNO2
STXBP2 — Leukocyte Migration
POLD4
CAPG
ARPC4
STX4
CYBA
GRN
CLDN4
AGPAT2
STXBP2
MVP
TOP3B
TYMP
SBNO2
CNOT3
ARHGEF1
BOP1 & HSF1 — Peripheral Nervous System
FGFR1
SCRIB
GAL
PTBP1
FGFR3
UPF1
NUP188
TRIP6
HSF1
BOP1
C20orf27
SLC52A2
EIF3B
SMARCA4
CCNF
BOP1 & HSF1 — Ubiquitin-dependent Protein Catabolic Process
EIF4G1
VARS
RAD23A
RNF126
SENP3
UBE2M
ARHGDIA
SMARCA4
POLRMT
HSF1
BOP1
APRT
ILF3
POLR2E
ATP5F1D
RBM42
DHX30
0.0
0.5
1.0

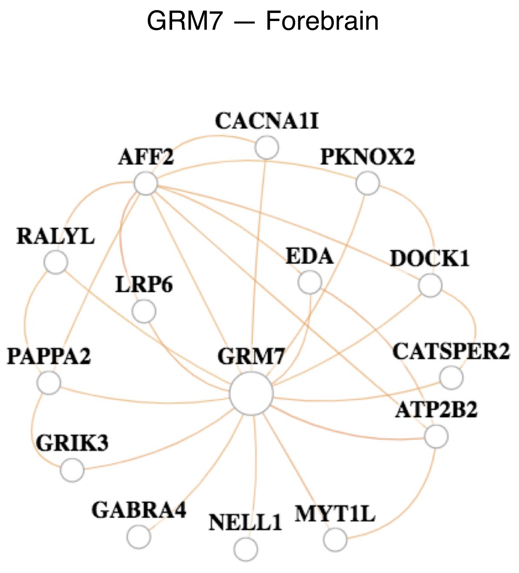


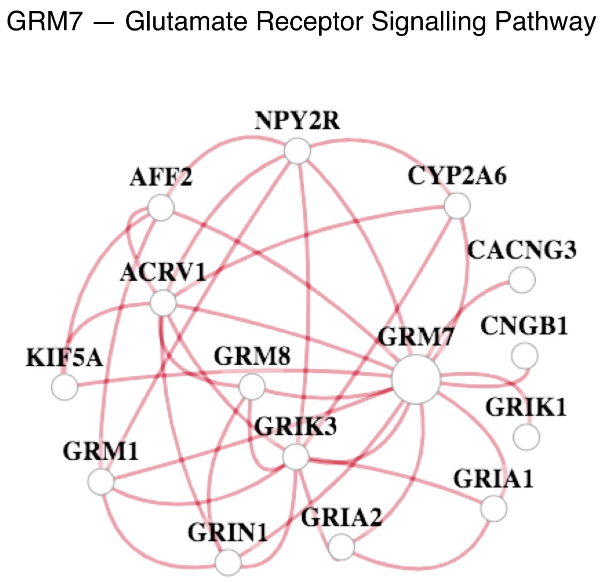


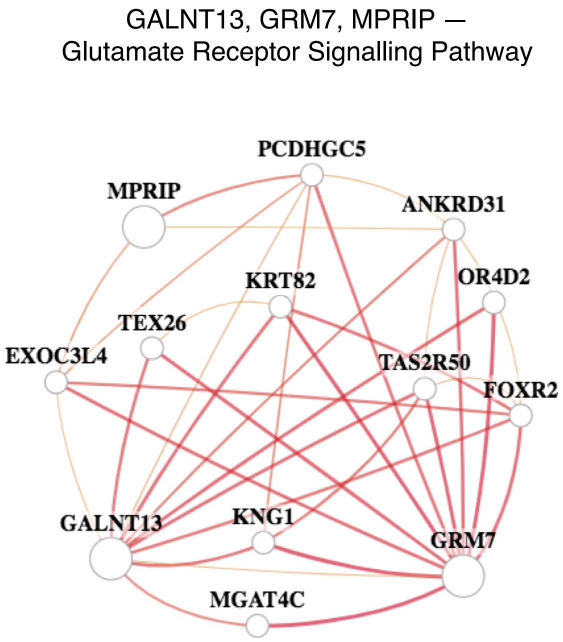


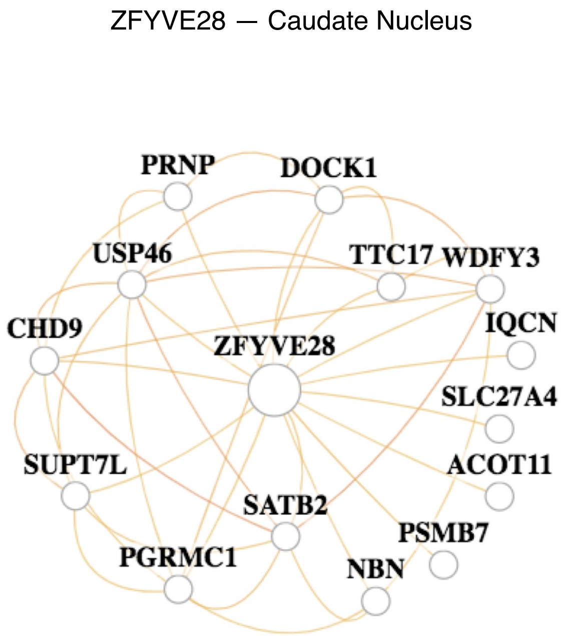


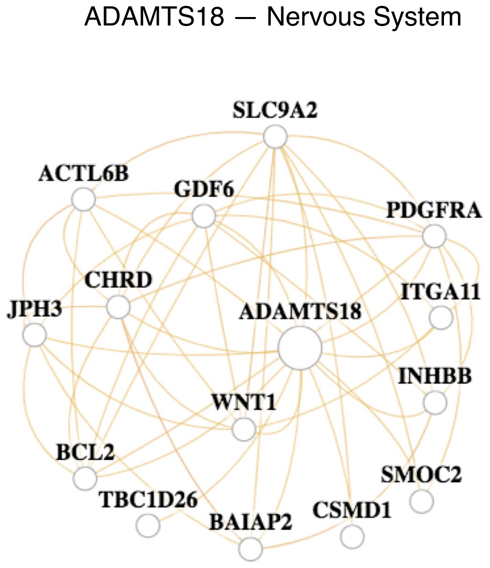


Supplementary Fig. S4. Predicted tissue- and biological process-specific gene-interaction networks for GALNT13, STXBP2, BOP1, HSF1, GRM7, MPRIP, ZFYVE28 and ADAMT18 in GIANT. The edges in a network indicate the predicted posterior probabilities of gene interactions between two genes. Top 14 genes highly connected to the query gene (large circle) are included in these networks.

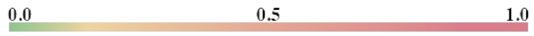

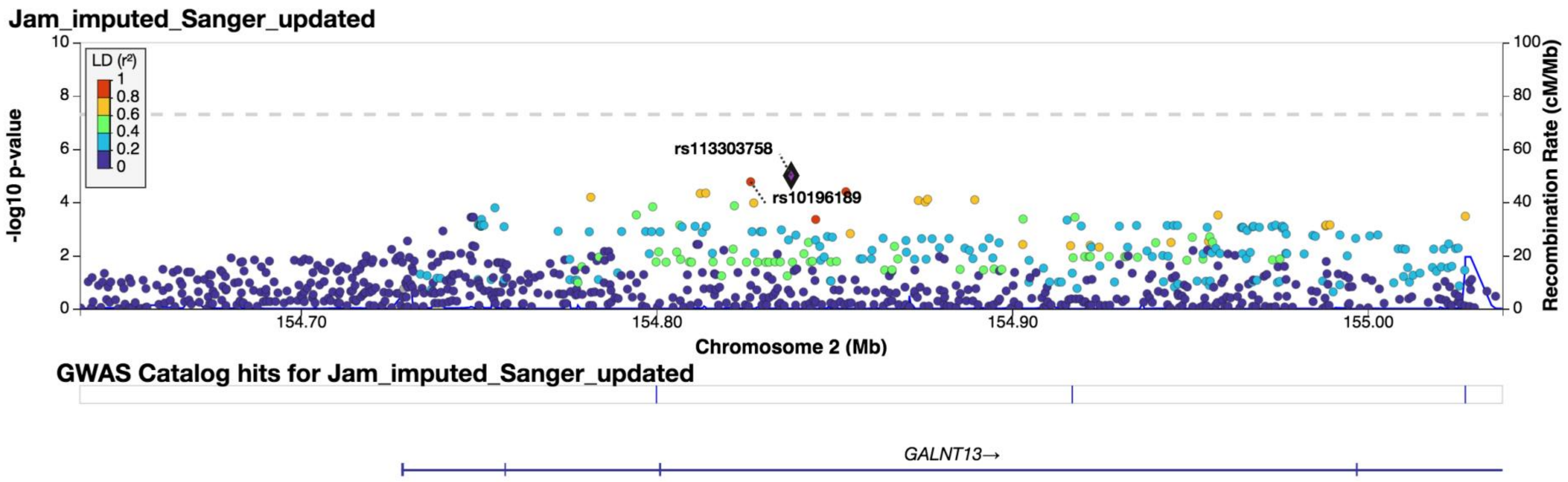


Supplementary Fig. S5. Regional association plot of 1Mb region surrounding the index SNP rs113303758 in *GALNT13* following Sanger Server imputation in the Jam cohort. -log10 transformed *P* value indicates the strength of the association with elite sprint performance. The level of LD and the recombination rate are estimated using the 1000 Genomes AFR population in LocusZoom v0.14.0.

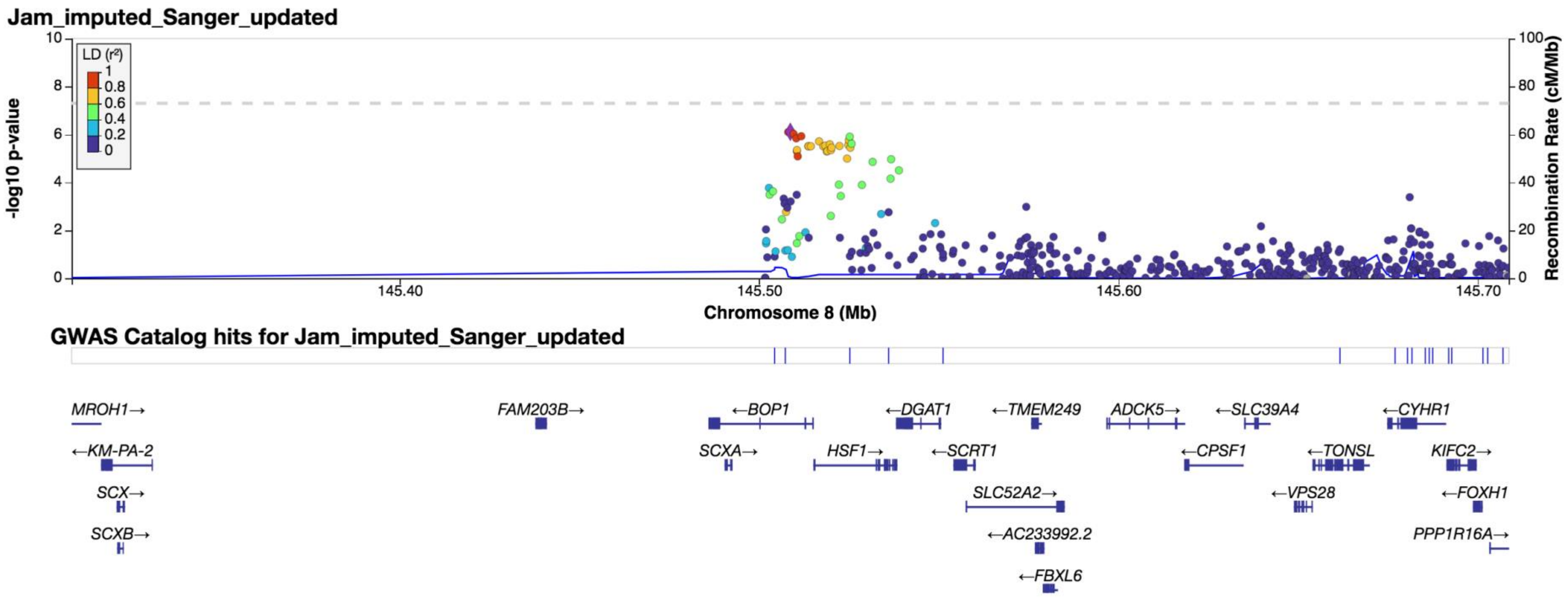


Supplementary Fig. S6. Regional association plot of 1Mb region surrounding the index SNP rs4977199 in *BOP1* following Sanger Server imputation in the Jam cohort. -log10 transformed *P* value indicates the strength of the association with elite sprint performance. The level of LD and the recombination rate are estimated using the 1000 Genomes AFR population in LocusZoom v0.14.0.

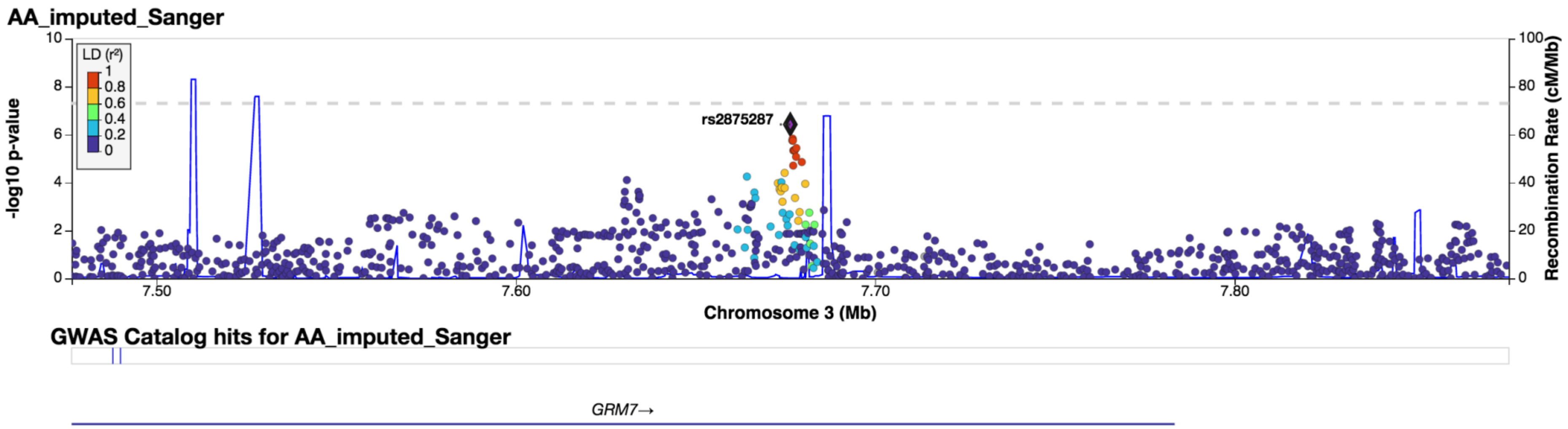


Supplementary Fig. S7. Regional association plot of 1Mb region surrounding the index SNP rs2875287 in *GRM7* following Sanger Server imputation in the A-A cohort. -log10 transformed *P* value indicates the strength of the association with elite sprint performance. The level of LD and the recombination rate are estimated using the 1000 Genomes AFR population in LocusZoom v0.14.0.

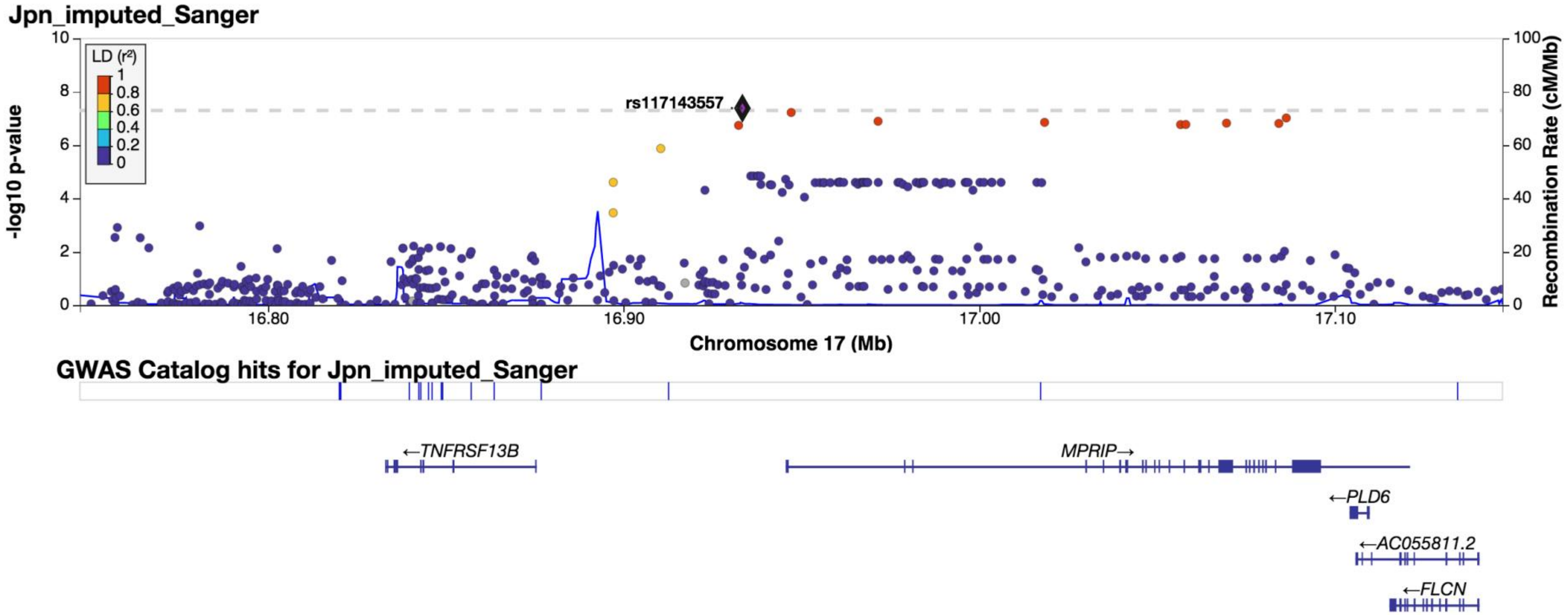


Supplementary Fig. S8. Regional association plot of 1Mb region surrounding the index SNP rs117143557 nearest *MPRIP* following Sanger Server imputation in the Jpn cohort. -log10 transformed *P* value indicates the strength of the association with elite sprint performance. The level of LD and the recombination rate are estimated using the 1000 Genomes AFR population in LocusZoom v0.14.0.

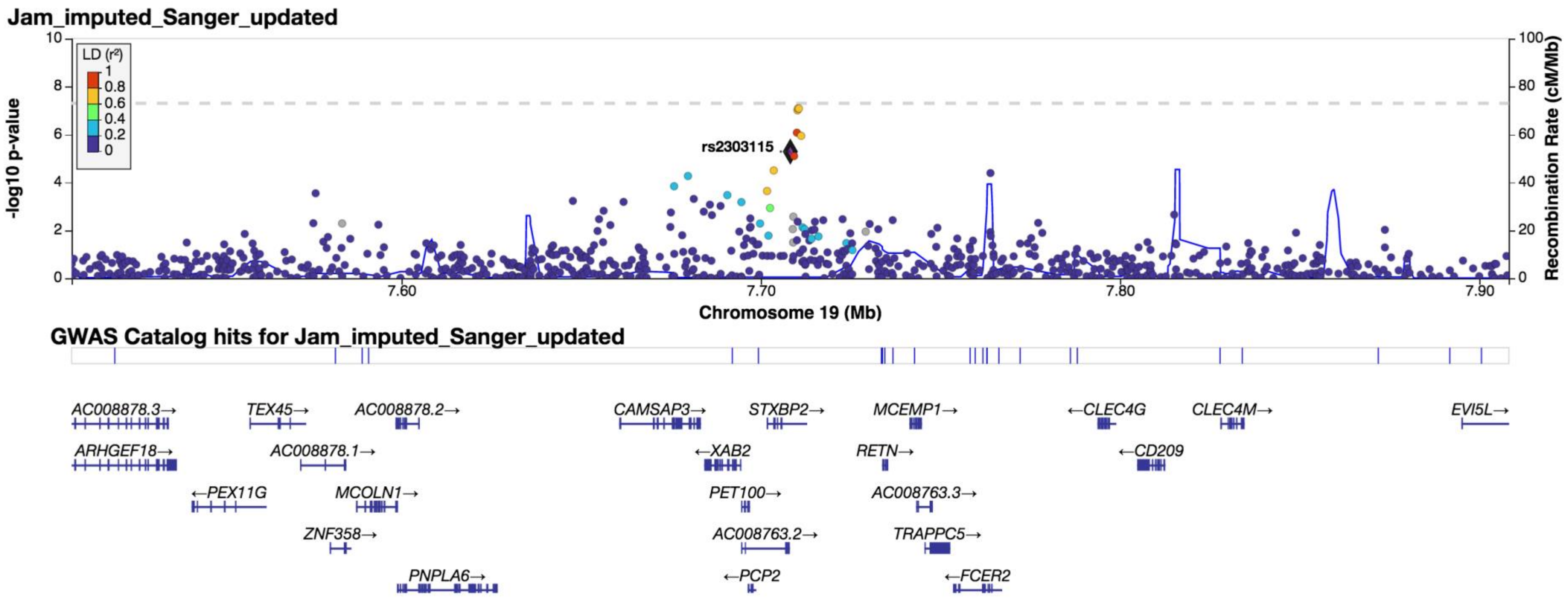


Supplementary Fig. S9. Regional association plot of 1Mb region surrounding the index SNP rs2303115 in *STXBP2* following Sanger Server imputation in the Jam cohort. -log10 transformed *P* value indicates the strength of the association with elite sprint performance. The level of LD and the recombination rate are estimated using the 1000 Genomes AFR population in LocusZoom v0.14.0.

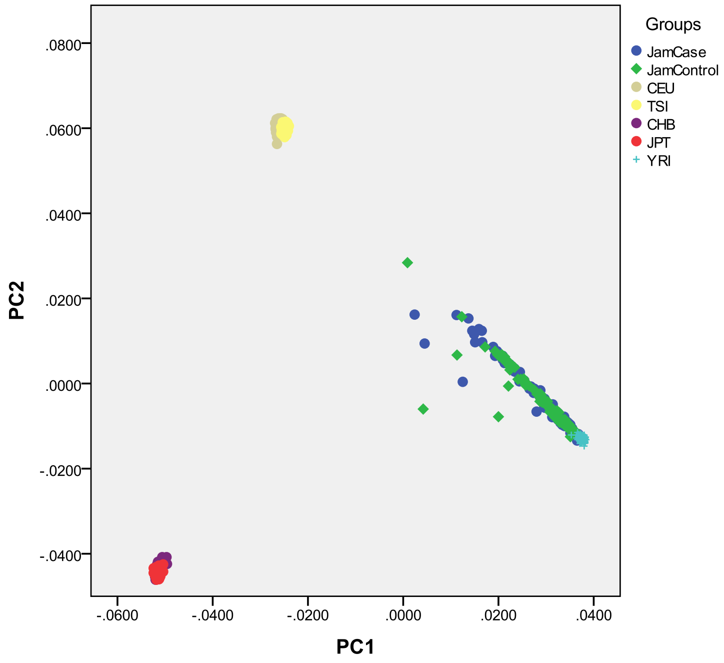


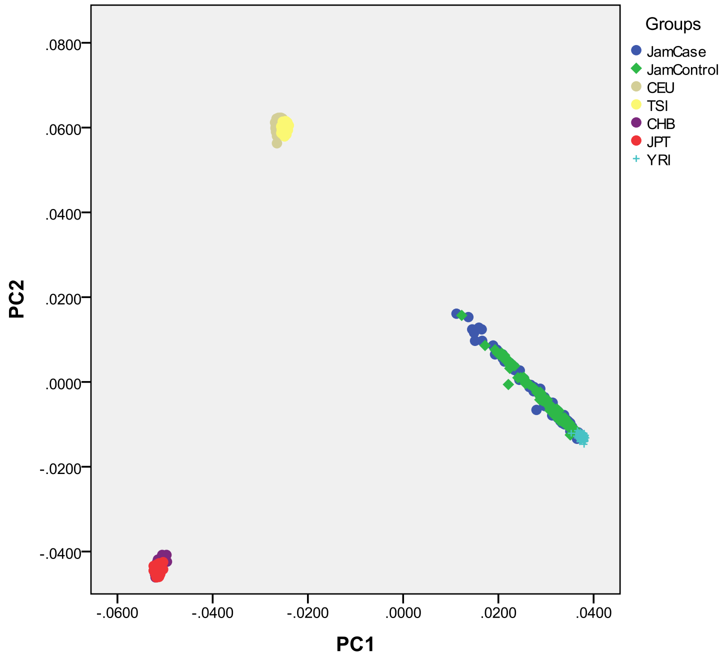


Supplementary Fig. S10. First two PCs for ancestry clustering of Jamaican sprint athletes and Jamaican controls alongside five Hapmap3 reference populations (CEU: Utah residents with Northern and Western European ancestry; TSI: Toscani in Italy; CHB: Han Chinese in Beijing, China; JPT: Japanese in Tokyo, Japan; YRI: Yoruba in Ibadan, Nigeria). Top: before the removal of outliers; bottom: after the removal of outliers.

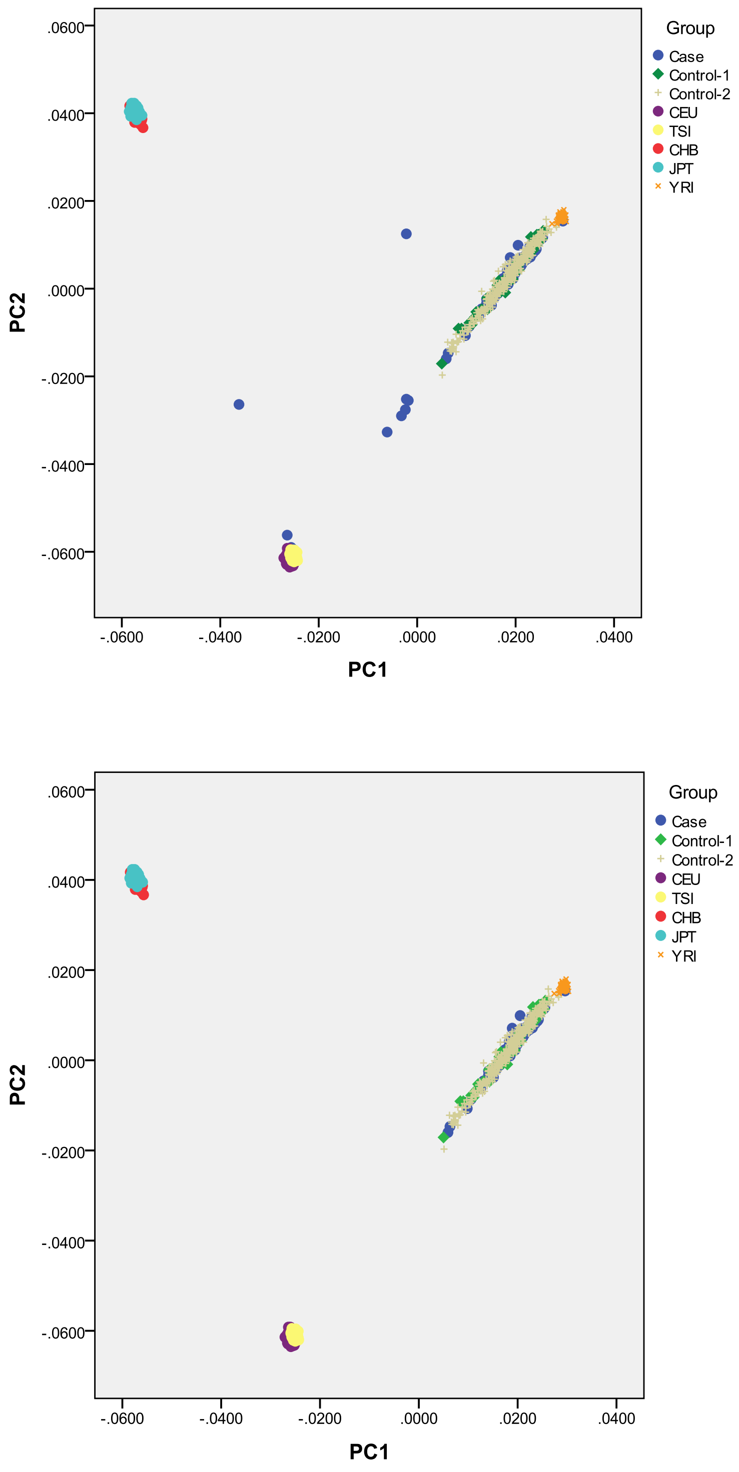


Supplementary Fig. S11. First two PCs for ancestry clustering of African-American sprint athletes and African-American controls alongside five Hapmap3 reference populations (CEU: Utah residents with Northern and Western European ancestry; TSI: Toscani in Italy; CHB: Han Chinese in Beijing, China; JPT: Japanese in Tokyo, Japan; YRI: Yoruba in Ibadan, Nigeria). Top graph: before the removal of outliers; bottom graph: after the removal of outliers. Control-1: 47 African-American controls; Control-2: 350 additional African-American controls.

Supplementary Table S1. Sample size of the two European replication cohorts for rs10196189 stratified according to geographical locations.

| | Population | Sprinters and jumpers | Other power/sprint oriented athletes | Endurance | Controls |
|---|---|---|---|---|---|
| **Cohort 1** | Belarusian | 35 | 53 | 16 | 205 |
| | Lithuanian | 41 | 37 | 181 | 392 |
| | Russian | 57 | 144 | 254 | 928 |
| | **Total** | **133** | **234** | **451** | **1525** |
| **Cohort 2** | Australian | 55 | 79 | 121 | 261 |
| | Belgium | 26 | 28 | - | 129 |
| | Greek | 54 | 13 | 23 | 55 |
| | Polish | 36 | 48 | 108 | 150 |
| | **Total** | **171** | **168** | **252** | **595** |

Note: No differences in allele frequencies or genotype distributions among the populations within each cohort in controls ($\chi^2$=9.32, *P*=0.16 in cohort 1; $\chi^2$=4.54, *P*=0.6 in cohort 2).

Supplementary Table S2. Genotype counts of rs10196189 stratified according to athlete types in each of the European replication cohorts.

| Cohort 1 | AA | AG | GG | Total |
|---|---|---|---|---|
| Sprinters (100-400m) and jumpers | 97 | 35 | 1 | 133 |
| Other power/sprint oriented athletes | 161 | 68 | 5 | 234 |
| Endurance athletes | 345 | 98 | 8 | 451 |
| Controls | 1190 | 315 | 20 | 1525 |

| Cohort 2 | AA | AG | GG | Total |
|---|---|---|---|---|
| Sprinters (100-400m) and jumpers | 115 | 48 | 8 | 171 |
| Other power/sprint oriented athletes | 123 | 43 | 2 | 168 |
| Endurance athletes | 200 | 46 | 6 | 252 |
| Controls | 439 | 148 | 8 | 595 |

Supplementary Table S3. Individuals failed sample QCs and excluded from further association analyses in the current GWAS cohorts.

| | **Jam** | **A-A** | **Jpn** |
|---|---|---|---|
| **Sex-check** | 2 athletes<br>2 controls | 6 athletes<br>2 controls | - |
| **Call rate** | 1 athlete<br>1 control | - | - |
| **Outliers of heterozygosity** | 4 controls | 6 athletes<br>2 controls | 2 controls |
| **Cryptic relatedness** | 1 athlete<br>4 controls | 2 athletes<br>1 control | - |
| **Outliers of PCA** | 3 athletes<br>4 controls | 13 athletes | - |
